# Individual longitudinal compliance to neglected tropical disease mass drug administration programmes, a systematic review

**DOI:** 10.1101/2022.10.06.22280483

**Authors:** Rosie Maddren, Santiago Rayment Gomez, Kathryn Forbes, Ben Collyer, Klodeta Kura, Roy Anderson

## Abstract

Repeated rounds of mass drug administration are the mainstay of transmission control for five of the 20 recognised neglected tropical diseases (NTDs); soil-transmitted helminths, schistosomiasis, lymphatic filariasis, onchocerciasis and trachoma. MDA programme efficiency is reliant upon participants swallowing treatment consistently at each treatment round, measured by the parameter compliance; the proportion of eligible participants swallowing treatment. Individually linked longitudinal compliance data is crucial for assessing MDA programmes, as such data will elucidate treatment behaviour patterns. Accurate monitoring of compliance across species will require the unanimous acceptance of epidemiological parameters across the research community. This review aims to update the review previously completed by Shuford et al (2016), which predominantly highlighted the interchange of parameters coverage (receiving treatment) and compliance (swallowing treatment). This review aims to find to collect the latest longitudinal compliance data reported by control programmes globally for the five MDA controllable species, searching PubMed and Web of Science in January 2022 for articles written since 2016 in English and Spanish. The review adhered to PRISMA guidelines and is registered with PROSPERO (registration number: CRD42022301991). Study title screening was aided by Rayyan, a machine learning software. Studies were considered for inclusion if primary compliance data for more than one time point, in a population larger than 100 participants were identified. All data analysis was conducted in R. A total of 89 studies were identified containing compliance data, 57 were longitudinal studies, 25 of which reported individually linked data which were analysed further. The association of increasing age with systematic treatment during was commonly reported. The review is limited by paucity of data. It is recommended for WHO to clearly define coverage, compliance, and longitudinal compliance in their treatment guides. Current definitions for species-specific guides contradict each other which may influence the incongruency seen definitions seen in this review.

**Author summary:** Neglected tropical diseases (NTDs) affect 1.74 billion people globally, often those in low socio-economic communities in tropical and sub-tropical climates. Five NTDs can be effectively treated using repeated administration of drugs across endemic communities, described as mass-drug administration (MDA). Repeated treatment is necessary due to re-infection of treated people by untreated people in these endemic communities. As such, increasing the number of people treated at each round is clearly critical to increase the number of parasite-free individuals, which will then latterly reduce the amount of re-infection to the community and therefore increase the chance of reaching elimination of transmission. Currently, the measurement of MDA success is focused upon coverage, the *acceptance* of treatment. However, not everyone who accepts treatment *swallows* the treatment, which arguably is more important to measure as it records the reality of the MDA success. This review aims to capture all the papers providing compliance data for soil-transmitted helminths, schistosomiasis, lymphatic filariasis, onchocerciasis and trachoma.

## Introduction

There are 1.74 billion people requiring interventions against neglected tropical diseases (NTDs). This has reduced by 600 million since 2010, yet considerable progress is still required to reduce the global burden of disease (1). The recently published WHO 2030 roadmap clearly demonstrates the link between socioeconomic disparity and NTD burden, with maps displaying GDP per capita and global NTD burden presenting two analogous images (1). NTDs are physically and financially devastating, and infection status (often resultant from poor hygiene or living conditions due to poverty) can further reduce an individual’s potential to earn, creating a downward spiral of well-being. As NTDs disproportionately affect individuals of low socioeconomic status, self-funded care is unfeasible for most affected, making global control of parasites predominantly reliant upon donations from pharmaceutical companies and operational support from international non-governmental organisations.

There are 20 NTDs recognised by WHO, five of which are responsible for the majority of the global NTD burden, namely lymphatic filariasis, onchocerciasis, schistosomiasis, soil-transmitted helminths (STH), and trachoma. These can by effectively treated with PC, significantly reducing morbidity. Entire endemic communities or at-risk subgroups receive mass drug administration (MDA) to treat the infection and control parasite transmission. Prior infection or treatment does not confer immunity to the parasite, thus repeated treatments are necessary due to re-infection of treated individuals by those who were not. These individuals who are not treated will continue to harbour parasites, and reinfection events will be common as transmission occurs through ingestion or contact with infectious larvae or parasite eggs found in human excreta, a pathway enabled in environments where sanitation is poor. Compliance (receiving and swallowing of drugs) across the targeted eligible population rarely, if ever, reaches 100%. This also results in the need for repeated treatments to lower parasite prevalence (2). The frequency and time interval between these repeated treatments is dependent upon prevalence levels and the life expectancy of the parasites’ infectious stage (2, 3). Since the increased efforts of NTD control inspired by the 2012 London Declaration (4), parasite prevalence levels have dropped sufficiently in some communities for elimination of transmission to be considered, thus requiring enhanced monitoring of current MDA activities and related parameters.

The success of MDA programmes should focus on the longitudinal patterns of treatment over time, rather than evaluating each MDA treatment round as a unique activity. Individual treatment history can be categorised as either non-treated (never treated), partially treated, or systematically treated. The longitudinal behaviour of these treatment categories can then be described as systematic (behaviour at round *x*, is dependent upon the behaviour at round *x*-1) or random (behaviour at time *x*, is independent upon behaviour at round *x*-1). Therefore, in administration units achieving high MDA coverage (receiving drugs) at each treatment round, if there is a high proportion of systematic non-treatment, i.e. the individual’s refusing treatment across each round are always the same, referred to as systematically non-treated individuals, then the targeted goal of parasite transmission elimination through reducing community reinfection will be improbable (5). The impact of individual treatment behaviour at each round of MDA delivery is currently unquantified in the literature, but has been suggested as a major hinderance to MDA programmes targeted with transmission elimination (6). Despite being rarely monitored, these longitudinal individual treatment behaviour patterns influenced by demographic, social and geographical factors will directly impact the number of future rounds of MDA required to elimination transmission of the targeted species (7). As such, before donor fatigue and economic pressures reduce the frequency and scale of NTD control programmes, improved monitoring of treatment behaviours to the currently available programmes using clearly defined parameters will be important to measure current successes accurately and comparably, and highlight areas required for intensified efforts to efficiently eliminate transmission of these parasites.

MDA delivery must achieve high compliance at each round for programmatic success. Currently, no guidelines exist, thus the recently published WHO 2030 NTD roadmap (1) would benefit from distinct coverage and compliance targets defined for each parasite species suitable for MDA of PC, as previously highlighted by Krentel *et al.* (2013) (8). As such, longitudinally monitoring of compliance is poorly actioned by the multiple control programmes across the NTD research landscape, as noted by a seminal paper on this topic by Shuford *et al.* (2016) (9). Limited analysis exists documenting age- and gender-stratified demographic factors of treatment behaviour. Elucidating any such trends in MDA behaviour will enhance the potential of future MDA rounds, for example the targeted dissemination of sensitisation activities to improve compliance in key groups.

Reasons for non-compliance at MDA are commonly reported, whereby MDA participants qualitatively report why they did not participate in MDA; largely divided into reasons for not being reached by a community drug distributor (CDD), or reasons for not swallowing drugs once reached by CDD. The importance this non-treated proportion of the population holds in the context of transmission elimination is dependent upon the R_0_ of the disease. If there is a high proportion of non-treatment within a population this is problematic for diseases with a higher R_0_ compared to those with a low R_0_ as there is more parasite transmission. Albeit a reductionist approach, the population requiring treatment can simply be thought of as R_0_-1/ R_0_, therefore for R_0_ = 2, 50% of the population need to be treated whilst R_0_ = 6, 83% of the population need to be treated (10, 11).

The interaction with CDD and participant can be divided into three key decision points of MDA behaviour, demonstrated in **Fig 1**. Similar decision trees have been published to clarify these parameters (12, 13). The three decision points define the epidemiological parameters, often used interchangeably to the detriment of monitoring and evaluation comparability of control programmes (9). For clarity, in this paper (and in keeping with that described by others (8, 9)), the term *coverage* refers to the proportion of the eligible population contacted by CDD, who received the offered drugs. *Compliance*, often confused or conflated with coverage, is referred to in this paper as the proportion of the eligible population who were contacted by CDDs and swallowed the received drugs. The importance of the distinction between these two epidemiological parameters in NTD research was first highlighted by Babu and Babu, 2014, in the context of reaching lymphatic filariasis transmission break in India (14), but has been identified in other disease areas since 1982 (11).

**Fig 1:**
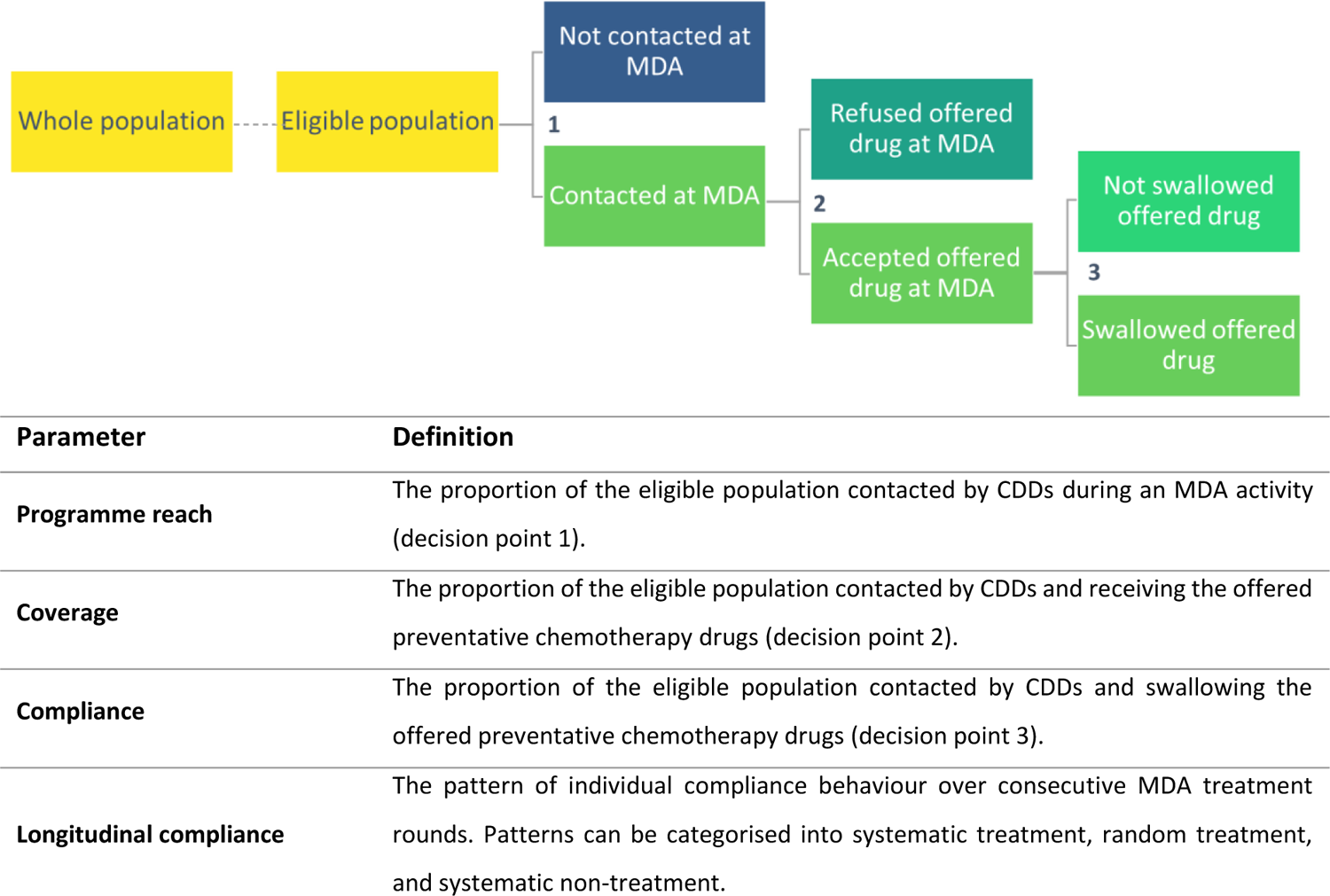
Decision tree of MDA treatment behaviour, showing the three behavioural decision points of MDA delivery and their definitions, according to these previously defined parameters by Shuford et al. (2016) (9). Abbreviations: CDD – community drug distributor.

Of crucial importance to control programmes, is the pattern, or lack thereof, of *longitudinal compliance* in individuals. The term longitudinal compliance is defined in this paper as the monitoring of individual treatment behaviour across multiple time points. Preferably, this would be measured by the gold standard of compliance measurement, directly observed treatment (DOT), avoiding any recall bias or desirability bias created from participant’s recalling past treatment. Longitudinal compliance is often misconstrued as *repeated cross-sectional compliance*, the repeated measurement of a random sample of the same population whereby individuals missing from different time-points are not removed. By chance, individuals may be sampled multiple times, but as the focus of the measurement at the population-level, not individual, the fine-scale granularity of individual-level data is diluted as the parameter measures a broader snapshot of compliance in time. From a longitudinal compliance dataset, individuals who systematically do or do not swallow drugs over consecutive rounds of MDA can be identified, referred to as *systematically treated* or *systematically non-treated*, respectively (6, 15). The terminology used to describe systematic non-treatment has evolved over recent years, to reflect the agency held by the participant and their behaviour such that systematic non-treatment has previously been referred to as non-adherence, or systematic non-compliance.

Coverage and compliance, if defined at all, are often confused in the literature. High coverage rates can occlude the realities of low drug ingestion (compliance), demonstrating the need for distinct measures. Incongruent denominators and numerators for both parameters are frequently reported, often leading to inappropriate calculations applied for each parameter, requiring careful interpretation of individual studies. Heterogeneity between the population reached, accepting treatment, or swallowing treatment over the eligible, at risk, or total population have all been employed, despite Shuford’s review published in 2016 presenting clear definitions for each parameter and work by Babu and Kar on lymphatic filariasis published in 2004 (16). The lack of consistently defined numerators and denominators for these parameters has been previously noted by Anderson *et al.* (2017) (17). Through deterministic models of parasite transmission and control, Anderson and team demonstrated the relationship between coverage and compliance upon MDA impact and the importance of monitoring these parameters in MDA programmes. Modelling systematic compliance patterns using deterministic and stochastic models, the group showed that, in a low helminth prevalence setting, populations displaying random treatment patterns over multiple rounds of MDA had a larger impact on prevalence decrease than populations displaying systematic non-treatment (18). This is directly relevant to programmes targeted with reaching transmission elimination, as these are inherently situations where low prevalence has been achieved such that a few persistent non-compliers to treatment can sustain transmission.

Improving compliance to treatment will not only increase the number of treated individuals, but these different longitudinal patterns of compliance within populations also influence the transmission dynamics of the parasite (6, 18) and thus is crucial to understanding the potential for transmission elimination in a defined population. Intuitively, a systematically non-treated population presents the greatest risk to control programmes compared to those who exhibit random patterns of treatment at each round of MDA. If a high proportion of the population are systematically non-treated (the same people refusing treatment each round), these individuals likely harbour a high proportion of the community parasite burden (parasite reservoir), consistently shedding parasite larvae or eggs into the local environment preventing the targeted goal of elimination of transmission. In areas of poor hygiene and sanitation, it is likely that this contamination from untreated individuals will re-infect the treated population, sustaining transmission (9). In randomly treated populations, individuals will, by definition, have typically received at least one round of treatment given reasonable levels of coverage (>75%), therefore reducing the likelihood that individuals harbour a reservoir of infection. This treatment pattern is therefore favourable, especially in the context of elimination (interruption of transmission), as is the current goal for onchocerciasis, or elimination as a public health problem (targeted for lymphatic filariasis, STH, SCH and trachoma) (1).

For helminth infections in particular, past infection does not confer strong acquired immunity hence, post treatment, all individuals are susceptible to repeated reinfection. Regardless of an individual’s previous treatment behaviour, they will become reinfected if they contact infectious eggs or larvae found in the environment. There is a small body of evidence to suggest low levels of immunity develops with age, opposing the argument of age-related exposure to infectious agents, however this factor is not sufficient to counter-act the force of re-infection (19). More substantial research exists to suggest immunity can be acquired for lymphatic filariasis, onchocerciasis, and trachoma infections. The concept of re-infection after treatment is especially important when control programmes cease MDA efforts after a targeted number of rounds or defined high level of coverage is achieved. Unless transmission is completely eliminated, infection in the community will continue from the infectious material surviving in the environment and harboured in untreated people, at a rate defined by the volume of infectious material in the environment - a concept referred to as ‘bounce-back’ (10). Only when compliance rates (and therefore by virtue, coverage rates) are high enough to reduce the probability of a female worm producing enough transmission stages that go on to survive to reproductive maturity in the human host, to less than unity in value (for all human helminths - STH, schistosomes, filarial worms, and onchocerca) will transmission cease (defined as “transmission break”).

Of interest in this paper is how compliance changes over time in populations: the significance of systematic treatment of individuals within a control programme. Historically, the description of demographic patterns, or lack thereof, of systematically non-treated populations over contiguous rounds of MDA has not been well-defined in the literature (9, 20). A systematic review of the available data published between the latest well-performed review by Shuford *et al.* (2016) (9) to 2022 was conducted to update the previous review and collect all available compliance data for the five treatable NTDs; STH, schistosomiasis, lymphatic filariasis, onchocerciasis and trachoma. If sufficient data is available, the review will elucidate if systematic non-treatment differed between age, gender, or other identified demographic subset of the population and if these treatment patterns are random or systematic in nature.

## Methods

The systematic review was conducted in line with the PRISMA guidelines (21) (**Supporting Information 1**) and is registered with PROSPERO (Registration number: CRD42022301991). Studies detailing systematic non-treatment to MDA for lymphatic filariasis, onchocerciasis, schistosomiasis, STH, and trachoma were searched using a Boolean strategy across PubMed and Web of Science databases in January 2022. The search strategy used and resulting PRISMA workflow are detailed in **Supporting Information 2 and 3**. The original review conducted by Shuford *et al.* (2016) (9) was considered to be completed comprehensively, therefore only studies published from 2016 up to 2022 written in English and Spanish were considered for inclusion. Studies were screened against minimal inclusion criteria (due to the paucity of relevant studies identified), which were: 1) primary compliance data reported from community or school-based control programmes targeting lymphatic filariasis, onchocerciasis, schistosomiasis, STH, and trachoma infections, 2) at least two defined time points of reported data, and 3) sufficiently large population sample (>100 participants). Where reported, reasons given for non-treatment were collected for subsequent meta-analysis. Care was taken to ensure that all data collected was explicitly defined as compliance (swallowing of drugs). Data described as parameters other than compliance, but using the correct compliance calculation were also recorded, with the variation of parameter definitions noted for later analysis. Nomenclature variations used to describe compliance included “programme / epidemiological coverage”, “uptake”, and “adherence”, as previously noted by Shuford *et al*. (2016) (9).

The 3649 studies generated from the three databases and associated searches were amalgamated in Excel, and de-duplicated based upon DOI and title matches. The 2529 remaining studies were then transferred to Rayyan, an online software using natural language processing and machine learning for data evaluation in a collaborative capacity (22). Using this software, whilst blinded, authors (RM, SGR) conducted title and abstract screening for inclusion, categorising paper themes as quantitative (considered for inclusion, (n = 164)), qualitative (n = 40), modelling (n = 45), or not relevant (n = 2280). Any conflicting cases were discussed and resolved on a case-by-case basis. Despite the exclusion of qualitative studies from this analysis, the themes presented in these studies were used to help interpret the resultant data analyses from the review. The 164 studies marked as quantitative for inclusion were read in full and assessed for eligibility for the final stage of data extraction. The references of which were screened to capture any further relevant data, generating one additional article for inclusion. From the 165 studies identified, 89 studies (datasets) were eligible for data extraction. Due to the paucity of papers containing longitudinal individual compliance data, longitudinal papers were further categorised in a tiered format as containing; individual compliance data (recorded by CDD), individual compliance data (self-reported), and repeated cross-sectional compliance data. Datapoints were extracted when the information presented satisfying the extraction criteria was available for detailed stratifications such as age, multiple regions and individual parasite species. Multiple datapoints were also extracted for cases when both coverage and compliance were reported, stratified by age-group, gender, location and, programme year of collection. Data extracted from the 89 studies included the study type, sample type including age and gender stratifications where available, number of studied participants, length of study, geographic identifiers, drugs provided, definitions of numerator and denominator for reported parameters, terminology used to describe parameters, and reasons for not complying with treatment. The 89 studies marked for inclusion were analysed in R (4.0.3 (2020-10-10) “Bunny-Wunnies Freak Out”), all figures were generated using ggplot2() unless specified.

## Results

A total of 165 studies met the inclusion criteria for data extraction (see **Supporting Information 4**). There were 46 papers discarded from the full-text review (including the Shuford *et al.* (2016) paper) namely due to seemingly relevant papers not reporting any compliance data, such as prevalence studies, or data presented in a format ineligible for extraction e.g. averages (n = 25), to reporting secondary data (n= 8), inability to access full-text data (n = 6), targeted treatment studies (n = 3), and reporting on sample sizes smaller than 100 (n = 2). There were 30 studies identified as reporting cross-sectional (n = 7) and longitudinal (n = 23) coverage data. Removing the irrelevant studies, and those reporting only coverage data, left a total of 89 studies to be considered in the analysis, a descriptive overview of which is provided in **Table 1**.

**Table 1:** Summary overview of the 89 studies included in the analysis split by the temporal format; either cross-sectional or longitudinal data. The full data extraction of the 165 studies is provided in **Supporting Information 4**. Frequency and percentage of studies falling into each category is shown as “n (%)”. Definitions: LF – lymphatic filariasis, N – number of studies, Oncho – onchocerciasis, preSAC – preschool-aged children, SAC – school-aged children, SCH – schistosomiasis, STH – soil-transmitted helminths.

|  |  | Cross-sectional<br>(N = 32) | Longitudinal<br>(N = 57) |
| --- | --- | --- | --- |
| **Sample Population** | preSAC | 0 (0) | 0 (0) |
|  | SAC | 8 (25) | 6 (11) |
|  | Adults | 2 (6) | 9 (16) |
|  | Community | 22 (69) | 38 (67) |
|  | Other | 0 (0) | 2 (4) |
| **Stratification** | Age | 12 (38) | 13 (23) |
|  | Gender | 13 (41) | 14 (25) |
|  | Region | 18 (56) | 22 (39) |
| **Species** | STH | 9 (28) | 9 (16) |
|  | SCH | 11 (34) | 6 (11) |
|  | Oncho | 4 (12) | 18 (32) |
|  | LF | 16 (50) | 29 (51) |
|  | Trachoma | 3 (9) | 3 (5) |

Across the 89 included studies, those stratifying compliance data by location were frequent, employed by 56% and 39% of the cross-sectional and longitudinal studies, respectively. Age and gender stratifications were reported by a smaller proportion of studies. Whole communities most frequently formed the sample population, followed by 69% and 67% of cross-sectional and longitudinal studies, respectively. Sample populations categorised as ‘other’ used non-standard age breakdowns such as 0-14 year-olds, and 2-14 year-olds. The mean (arithmetic) sample size of cross-sectional studies (x^-^ = 4965) was considerably smaller than that of longitudinal studies (x^-^ = 1,257,386). Five of the included longitudinal studies reported compliance in populations ranging from 1-400 million, covering several districts during national control programmes in Myanmar (23), China (24), Sierra Leone (25, 26), and the Dominican Republic (27).

Within the 89 studies included in this analysis, 78 studies reported MDA data for a single parasite species. An additional 30 datapoints were generated by 11 studies reporting compliance data for two (n = 10), three (n = 12), and four (n = 8) species together. The total 108 species-specific datapoints from the 89 datasets are shown in **Fig 2**. Overall, 45, 22, 18, 17 and 6 studies reported compliance data for lymphatic filariasis, onchocerciasis, STH, schistosomiasis, and trachoma. Studies focusing solely upon lymphatic filariasis (n = 37) and onchocerciasis (n = 17) were most numerate and were found to be most consistently published between 2016 and 2022 (depicted as orange and yellow points, respectively, in **Fig 2**). Papers solely focused on STH were less so (n = 9) but were the most frequently reported species (n = 3) in combination with schistosomiasis. Trachoma studies were least common (n = 6) and were reported only as a single species, never in combination with other parasites presumably due to different control methods. No trachoma compliance studies have been published since 2019.

**Fig 2:**
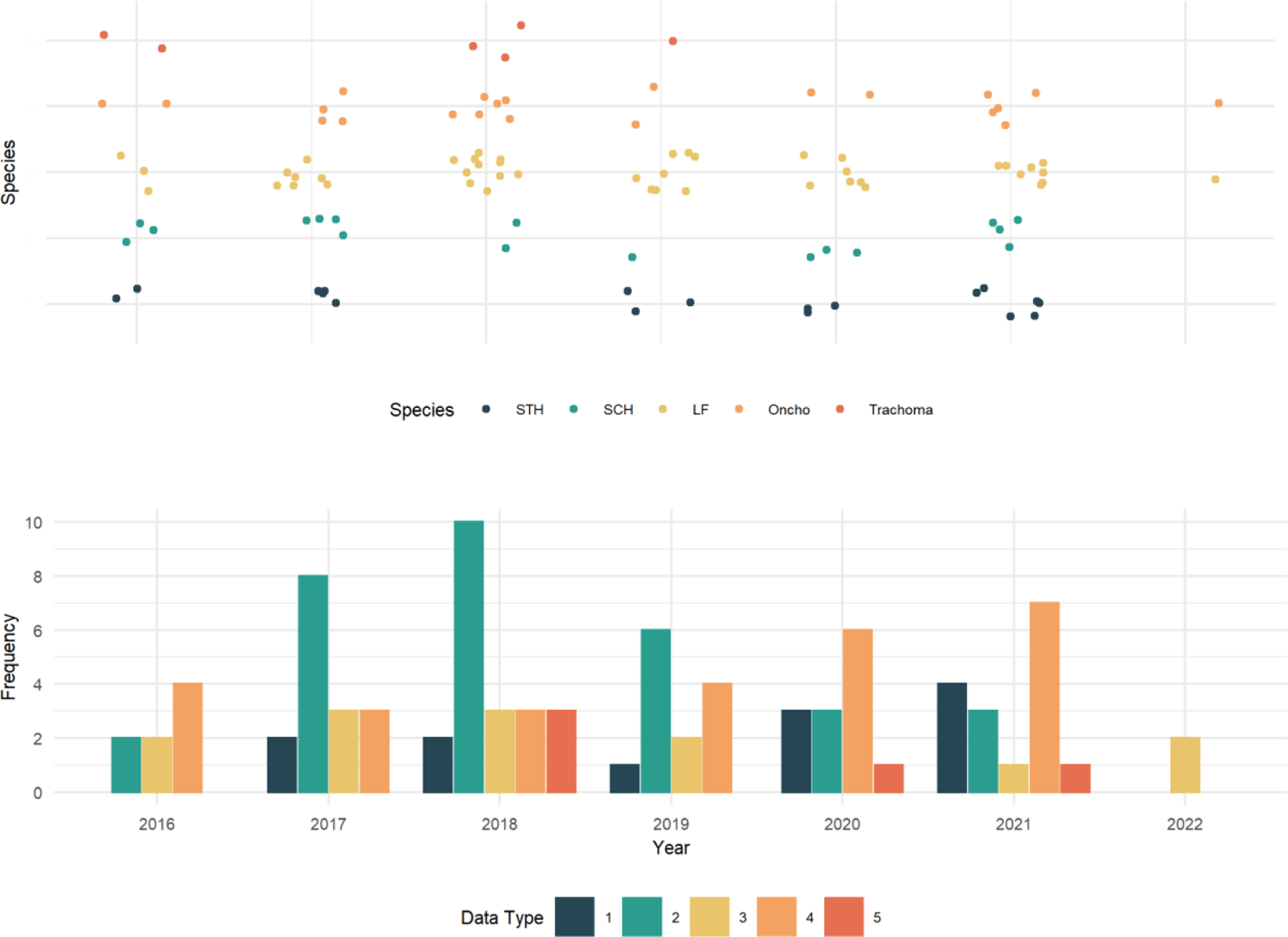
Number of studies published each year for each species, from 2016 to 2022. 11 studies are represented as 30 datapoints where studies reported data for more than one species, totalling 96 datapoints from 89 studies. Points are coloured by each of the five reported species. Point shape is defined by the type of data presented, ranked in order of quality: 1 – individual longitudinal compliance, 2 – both longitudinal and cross-sectional compliance, 3 - repeated cross-sectional compliance, 4 - cross-sectional compliance, 5 – undefined cross-sectional. Definitions: LF – lymphatic filariasis, Oncho – onchocerciasis, SCH – schistosomiasis, STH – soil-transmitted helminths.

From the 89 studies, an additional 11 geographical datapoints were generated from four studies reporting data from multiple countries, providing a total of 96 geographic datapoints. Globally, studies reporting compliance from Tanzania (n = 11) and Ethiopia (n = 9) were most prevalent, as depicted in **Fig 3**.

**Fig 3:**
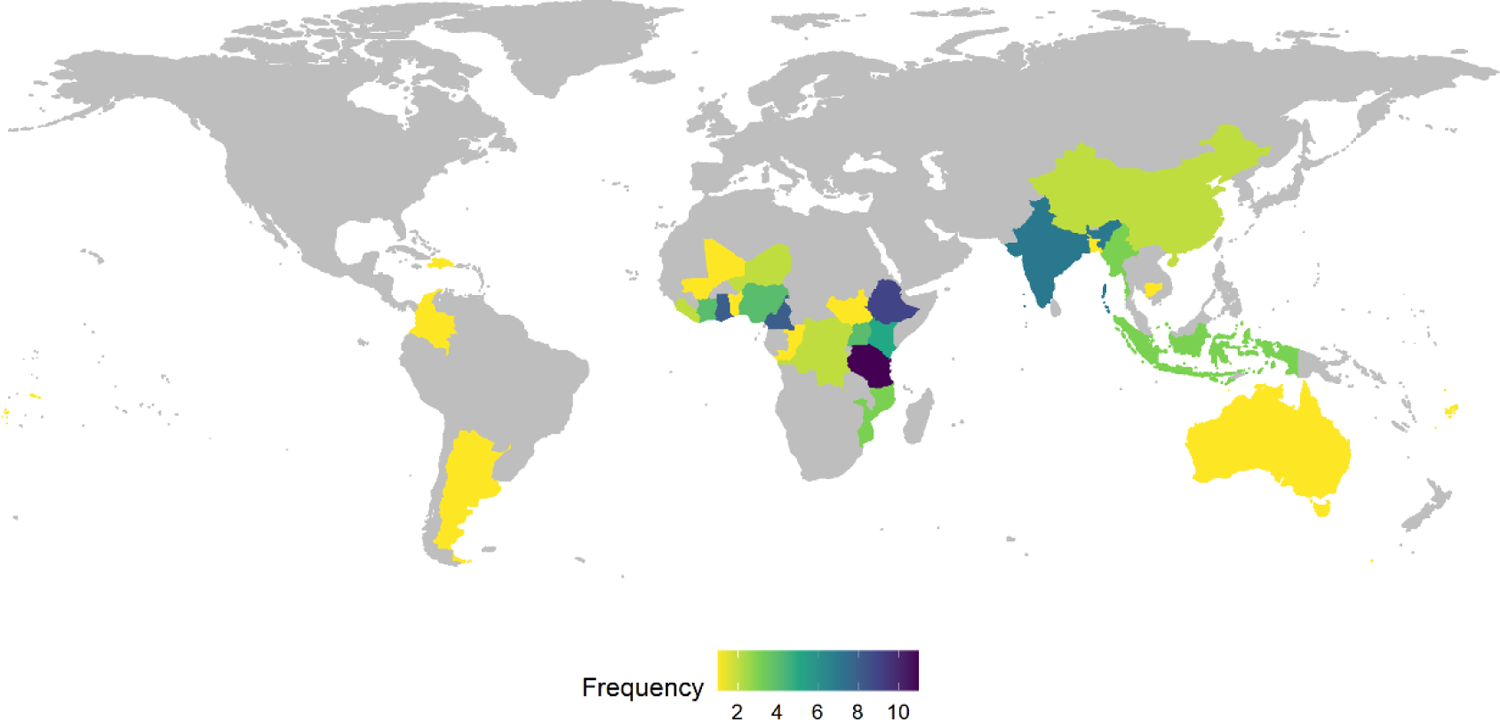
Choropleth map of studies published globally from 2016 to 2022. Countries where no papers were published are shown in grey. Four studies are represented as 11 datapoints where studies reported data for more than one country, totalling 108 datapoints from 89 studies.

Due to the paucity of available literature detailing individual longitudinal compliance, a hierarchy of studies was created, encompassing both longitudinal and cross-sectional studies as detailed in **Table 2**. Often, compliance was misconstrued as coverage, and the terminology, numerator, and denominators used to describe these parameters were heterogeneous, as previously noted by Shuford *et al.* (2016) (9). The reported numerator, denominator, and definitional calculations were carefully analysed for all studies where provided, and the correct parameter was assigned to the data where necessary.

**Table 2:** Data type and definition used to classify compliance papers included in the analysis. Longitudinal compliance data was reported by both longitudinal studies (cohort, randomised controlled trials) and cross-sectional studies.

| Compliance Data Type | Study Type | Description |
| --- | --- | --- |
| **Individual – annually surveyed (n = 3)** | Cohort, randomised controlled trial | Longitudinal, individually linked, compliance data across >2 time points. Recorded during annual surveys either by CDD (directly observed treatment), or self-reported by participant. Focus: individual level. |
| **Individual – self-reported (time-specific or overall) summary (n = 22)** | Cross-sectional | Longitudinal compliance data over defined/undefined time-period. Self-reported compliance by an individual at one time-point, looking back in time e.g., self-reported compliance to 5 available MDA rounds. Focus: individual level. |
| **Repeated cross-sectional (n = 32)** | Cross-sectional | Compliance data at each round of MDA for a set cohort (e.g., village or district), repeatedly surveyed over several time points population repeatedly sampled. Individuals not consistently present across all time points are not removed. Focus: population level. |
| **Cross-sectional (n = 32)** | Cross-sectional | Compliance data for a single round of MDA. Focus: population level. |

Of the 89 studies, 57 (64%) contained longitudinal data which were further categorised as individual data (recorded by data collectors or self-reported), repeated cross-sectional data, or cross-sectional data (**Table 2**). The length of studies ranged from repeated cross-sectional compliance from 26 annual rounds of MDA in China (24), to repeated cross-sectional compliance from two biannual rounds in South Sudan (28) and Nigeria (29). There were 30 studies (36%) detailing cross-sectional (single time-point) compliance data, which were further categorised by the numerator and denominator provided. Studies not explicitly defining either of these values were considered “assumed”.

The compliance data type reported did not change significantly over time, as shown in **Fig 2**. Repeated cross-sectional studies were the most frequent study type, reported in 32 of the 89 studies, preceded by cross-sectional compliance (n = 27), both longitudinal and cross-sectional compliance (n = 13), individual longitudinal compliance (n = 12), and undefined cross-sectional compliance (n = 5). Studies reporting individual longitudinal compliance did slightly increase over time (navy bar), with four studies reported in 2021. Similarly, the number of cross-sectional compliance studies published also slightly increased (orange bar), with seven studies reported in 2021.

### Incongruent parameter definitions

Not all studies provided clear definitions of the parameter reporting compliance. Analysing the numerators and denominators provided, just 26 (29%) studies could be confidently identified as containing either cross-sectional or longitudinal compliance data; the proportion of the eligible population who were contacted by CDDs and swallowed the offered drugs. Compliance data defining either the correct numerator or denominator for compliance, not both, were categorised as assumed (n = 12). Due to the wide variety of definitions used for the same parameter across studies (see **Fig 5**), studies that did not define either numerator or denominator for the parameter given were classified as undefined (n = 5). These parameters were “epidemiological coverage”, “MDA participation”, “treatment coverage”, “MDA coverage” and “compliance”.

The parameter “treatment coverage” (n = 4) was used most frequently to describe cross-sectional compliance, whereas longitudinal compliance was most frequently, correctly referred to as “compliance” (n = 8). Compliance was only correctly used in three of the 32 cross-sectional studies, with the numerator and denominator clearly defined (13,30,31). The variety of parameters provided to describe cross-sectional and longitudinal compliance as defined in this review is shown in **Fig 5**.

### Individual longitudinal compliance data

Longitudinally following cohorts to identify individual patterns of treatment behaviour, and the frequency of each pattern in the population is essential, as previously described. The 57 longitudinal compliance studies were further broken down into three hierarchical categories as detailed previously in **Table 2 (rows 1-3)**, into individually-linked compliance data recorded at each round (the category originally desired for the review due to the data importance for MDA monitoring and evaluation), compliance data recorded retrospectively (individuals recalling treatment compliance over number of rounds or ‘ever’ complying to MDA rounds), or repeated cross-sectional compliance data. A sample of the extracted columns from the review is shown below for the 12 studies containing both annually surveyed and self-reported individual compliance data in **Table 3**.

**Table 3:**
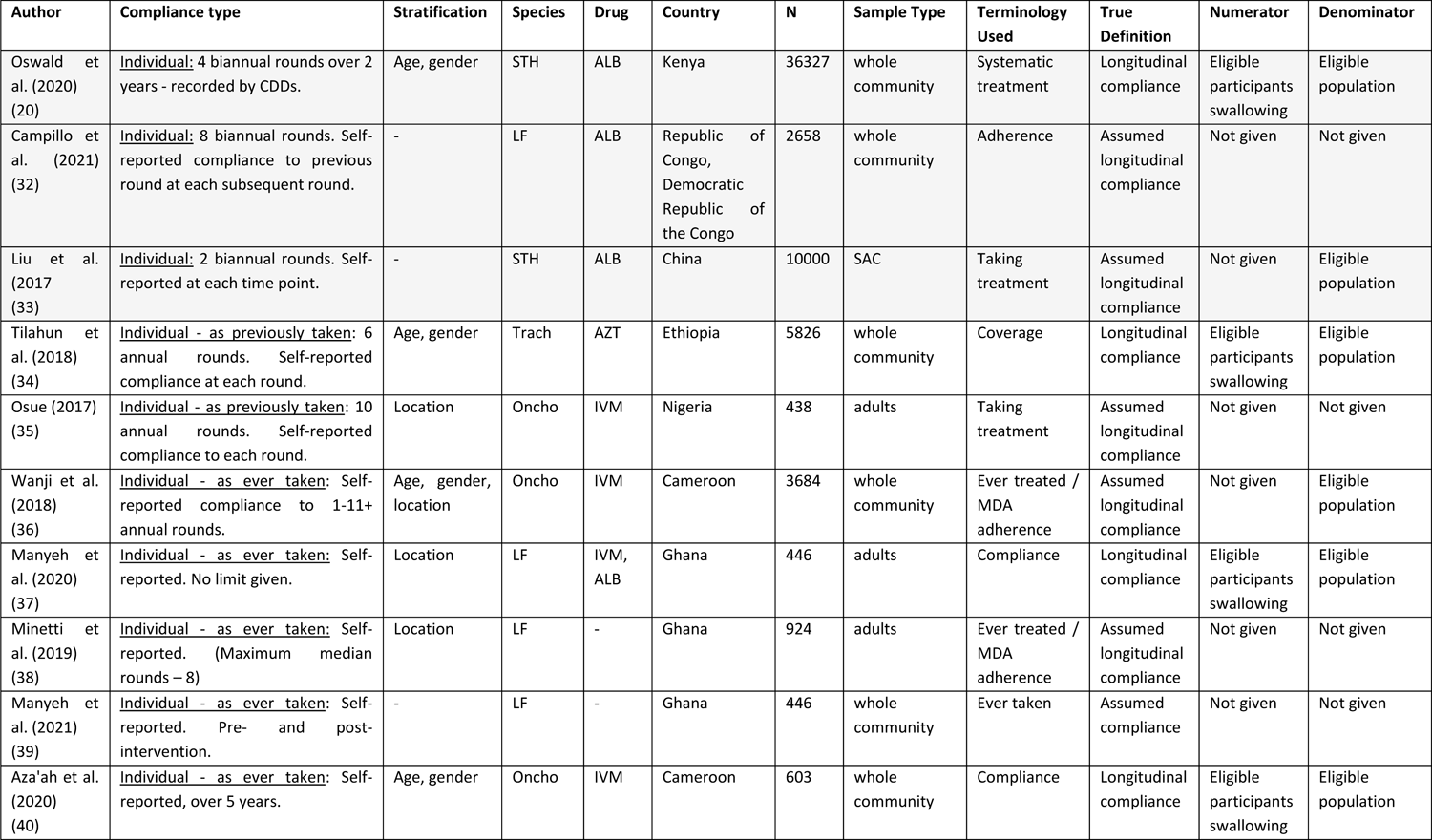
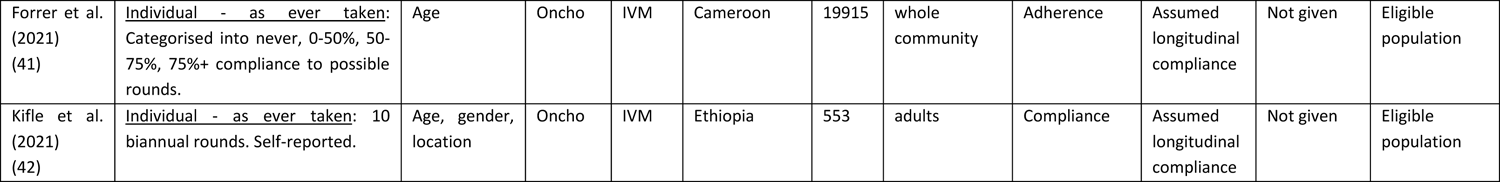
A selection of the extracted columns for the 12 studies satisfying the criteria for individual-level data – both annually surveyed by CDD or self-reported by participants (grey shading), and self-reported during a cross-sectional analysis. Available stratifications of compliance data are shown from the possible categories of age, gender, or location. The species targeted for control by MDA, and the drug offered for which compliance to is measured, is also given. The country, and the type and number of participants is also provided. The terminology used to describe compliance is shown, along with the ‘true’ definition according to this review given the numerator and denominator recorded (where given). Where there are incomplete numerator or denominators provided, compliance can only be assumed given the large heterogeneity of calculations used for the parameter. Abbreviations: ALB – Albendazole, IVM – Ivermectin, LF – lymphatic filariasis, Oncho – onchocerciasis, SAC – school-aged children, SCH – schistosomiasis, STH – soil-transmitted helminths, Trach – trachoma.

| Author | Compliance type | Stratification | Species | Drug | Country | N | Sample Type | Terminology Used | True Definition | Numerator | Denominator |
| --- | --- | --- | --- | --- | --- | --- | --- | --- | --- | --- | --- |
| Oswald et al. (2020) (20) | <u>Individual</u>: 4 biannual rounds over 2 years - recorded by CDDs. | Age, gender | STH | ALB | Kenya | 36327 | whole community | Systematic treatment | Longitudinal compliance | Eligible participants swallowing | Eligible population |
| Campillo et al. (2021) (32) | <u>Individual</u>: 8 biannual rounds. Self-reported compliance to previous round at each subsequent round. | - | LF | ALB | Republic of Congo, Democratic Republic of the Congo | 2658 | whole community | Adherence | Assumed longitudinal compliance | Not given | Not given |
| Liu et al. (2017) (33) | <u>Individual</u>: 2 biannual rounds. Self-reported at each time point. | - | STH | ALB | China | 10000 | SAC | Taking treatment | Assumed longitudinal compliance | Not given | Eligible population |
| Tilahun et al. (2018) (34) | <u>Individual - as previously taken</u>: 6 annual rounds. Self-reported compliance at each round. | Age, gender | Trach | AZT | Ethiopia | 5826 | whole community | Coverage | Longitudinal compliance | Eligible participants swallowing | Eligible population |
| Osue (2017) (35) | <u>Individual - as previously taken</u>: 10 annual rounds. Self-reported compliance to each round. | Location | Oncho | IVM | Nigeria | 438 | adults | Taking treatment | Assumed longitudinal compliance | Not given | Not given |
| Wanji et al. (2018) (36) | <u>Individual - as ever taken</u>: Self-reported compliance to 1-11+ annual rounds. | Age, gender, location | Oncho | IVM | Cameroon | 3684 | whole community | Ever treated / MDA adherence | Assumed longitudinal compliance | Not given | Eligible population |
| Manyeh et al. (2020) (37) | <u>Individual - as ever taken</u>: Self-reported. No limit given. | Location | LF | IVM, ALB | Ghana | 446 | adults | Compliance | Longitudinal compliance | Eligible participants swallowing | Eligible population |
| Minetti et al. (2019) (38) | <u>Individual - as ever taken</u>: Self-reported. (Maximum median rounds – 8) | Location | LF | - | Ghana | 924 | adults | Ever treated / MDA adherence | Assumed longitudinal compliance | Not given | Not given |
| Manyeh et al. (2021) (39) | <u>Individual - as ever taken</u>: Self-reported. Pre- and post-intervention. | - | LF | - | Ghana | 446 | whole community | Ever taken | Assumed compliance | Not given | Not given |
| Aza'ah et al. (2020) (40) | <u>Individual - as ever taken</u>: Self-reported, over 5 years. | Age, gender | Oncho | IVM | Cameroon | 603 | whole community | Compliance | Longitudinal compliance | Eligible participants swallowing | Eligible population |
| Forrer et al. (2021) (41) | <u>Individual - as ever taken:</u><br>Categorised into never, 0-50%, 50-75%, 75%+ compliance to possible rounds. | Age | Oncho | IVM | Cameroon | 19915 | whole community | Adherence | Assumed longitudinal compliance | Not given | Eligible population |
| Kifle et al. (2021) (42) | <u>Individual - as ever taken:</u> 10 biannual rounds. Self-reported. | Age, gender, location | Oncho | IVM | Ethiopia | 553 | adults | Compliance | Assumed longitudinal compliance | Not given | Eligible population |

A single study was identified as containing annual longitudinal compliance data, referring to the parameter as “systematic treatment”. Treatment behaviour of 36,000 individuals’ for four rounds of biannual MDA data for STH was recorded by CDDs at point of delivery, digitised, and individually-linked as part of the TUMIKIA project in Kenya (20). A further two studies contained annual assumed longitudinal compliance data, due to incomplete definitions of parameter numerators and denominators, referring to longitudinal compliance as either “treatment adherence” (32), or “taking treatment” (33). In both studies, individuals self-reported their compliance to the previously administered round of MDA, providing data for each round of lymphatic filariasis MDA in the Democratic Republic of the Congo and the Republic of Congo (32), and STH MDA in China (33).

Time-specific and overall longitudinal compliance studies were more common (n = 9). Two longitudinal studies asked respondents to recall their previous treatment behaviour at each round, over six annual rounds in Ethiopia for trachoma (34), and over 10 annual rounds in Nigeria for onchocerciasis (35). Three cross-sectional studies asked participants to recall their treatment behaviour during MDA for onchocerciasis over a specific time period, either five annual rounds (40) and 11+ annual rounds (36) in Cameroon, or 10 biannual rounds in Ethiopia (42). Two of these studies referred to compliance in line with the definitions considered for this review (40, 42). A further two cross-sectional studies asked participants to recall ever being treated during MDA for lymphatic filariasis in Ghana (37, 38). One study reported longitudinal compliance as a percentage of the possible rounds per participant based upon eligibility, breaking compliance into ‘never’, or complying with “≤50%”, “50-75%”, “≥75%” of possible rounds of MDA against onchocerciasis in Cameroon (41). One study reported the change in compliance before and after an interventional effort to increase MDA uptake for lymphatic filariasis control in Ghana (39).

A combination of cross-sectional and longitudinal compliance was reported by 13 studies, whereby compliance to a defined (43–50) or undefined (51–55) number of rounds were reported, alongside compliance at the latest round of MDA. Four of these studies defined compliance in line with the definitions considered for this review (45,51–53). Setting a high reporting standard, Osei *et al.* (2022) (53), Oswald *et al.* (2016) (20) and Krentel *et al.* (2016) (55) provided comprehensive demographic and descriptive factors and associations with receiving, and swallowing MDA through bivariate and multivariate logistic regression analysis. This high level of descriptive detail for coverage and compliance was rarely observed in the literature. The research landscape of MDA reporting and its impact on infection levels would greatly benefit from more studies following these detailed study design principles.

A total of 32 studies reported repeated cross-sectional compliance data over time periods ranging from a year of biannual treatment to 26 years of annual treatment. As these datasets follow compliance at a population level, this is not considered as individual longitudinal compliance.

### Study duration of longitudinal studies

From the 25 longitudinal studies, the majority consisted of studies asking participants how many rounds of MDA they swallowed treatment, over their lifetime or a defined period as shown in yellow in **Fig 4**. Annual and biannal studies were found at equal frequencies in the review (n = 5). Biannual stduies tended to follow fewer rounds of MDA, compared to annual studies.

**Fig 4:**
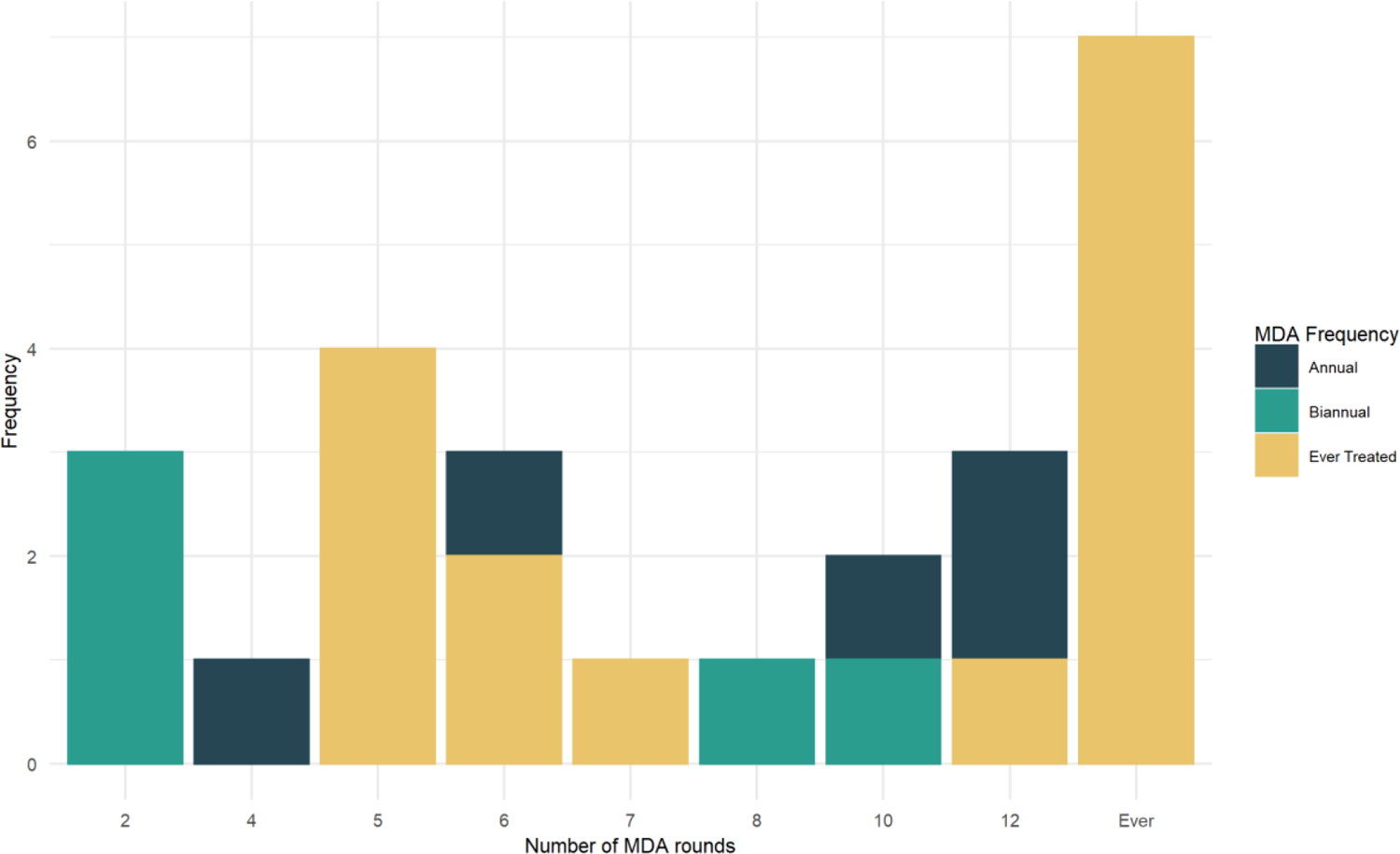
Length of study duration of longitudinal compliance studies by (n = 25). Bars are coloured by the frequency of rounds (annual, biannual) or the time frame in which participants were “ever treated”.

### Compliance versus coverage

From the 32 and 57 cross-sectional and longitudinal studies reporting compliance, 41% (n = 13) and 14% (n = 8) reported coverage data alongside compliance data. These 21 studies produced 173 datapoints of comparable coverage and compliance data for different demographic strata, namely age, gender, location, and programme year. Uniquely, Chami et al. (2017) (13) calculated the average marginal effects (AME) for coverage and compliance against a variety of factors. Most commonly, coverage was reported as a proportion of the eligible population (studies = 16, datapoints = 77), but studies also reported the parameter as a raw number (studies = 2, datapoints = 11) or as different named parameters such as epidemiological, programme and geographic coverage (studies = 3, datapoints = 85). There was a mean average difference of 4.91% (min: 0%, max: 30.8%) between coverage and compliance figures reported as a proportions. Similarly, there was a 11.9% (min: 7.04%, max: 21.5%) mean average percentage change between the raw data figures reported. A larger discrepancy of 17.7% (min: 10.3%, max: 45.9%) was noted between epidemiological / programme coverage, and compliance / geographical coverage. The difference between the two parameters is shown in **Fig 5**.

**Fig 5:**
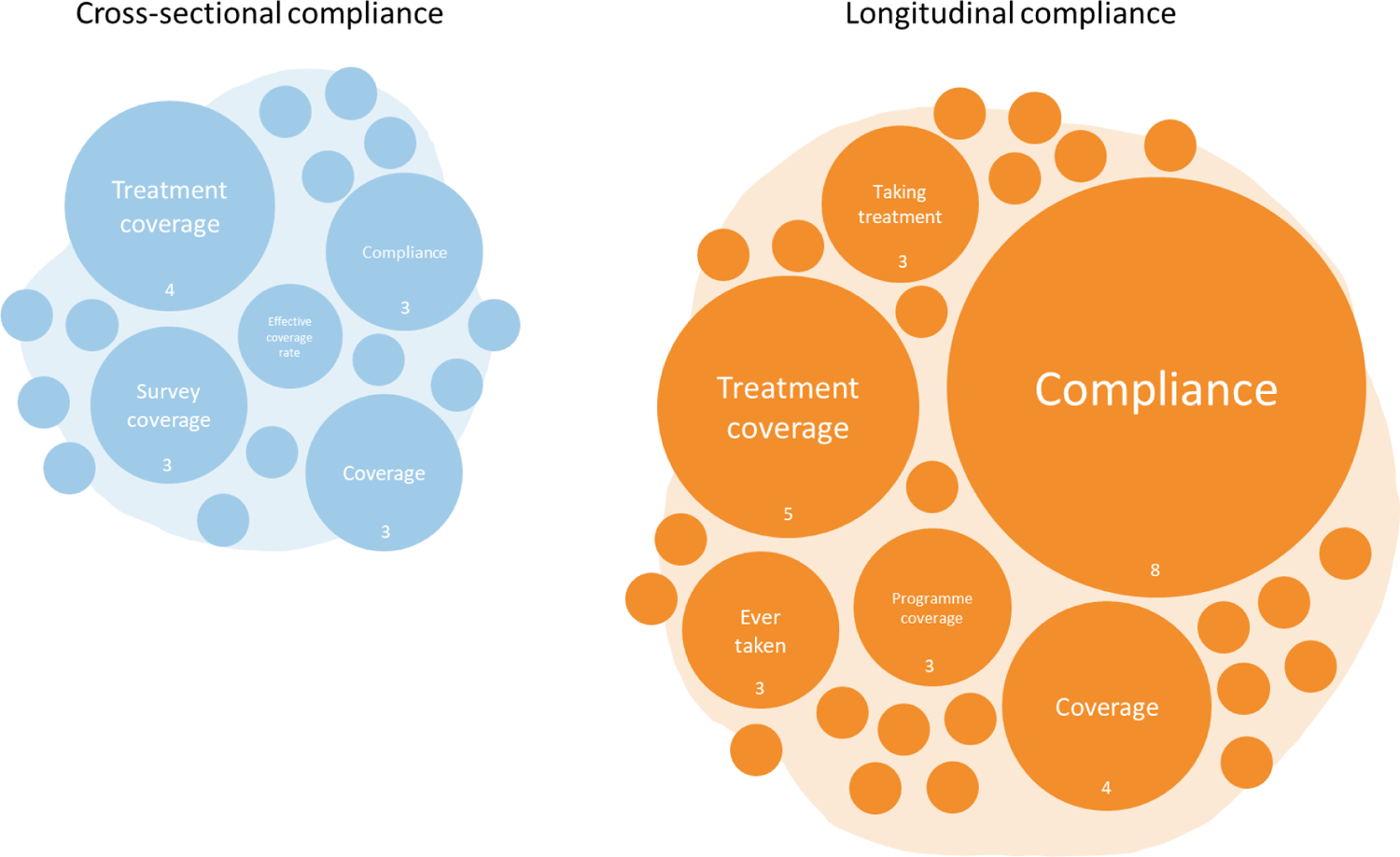
The difference between coverage and compliance figures reported by 21 studies as either proportions (**A**), raw data (**B**) or other parameters (**C**). Datapoints are coloured by the study type, either cross-sectional (13 studies, 69 datapoints) or longitudinal (8 studies, 104 datapoints). An average trendline of the reported data is shown in solid grey. The shaded line represents the conditional mean function using a linear model calculated by the geom_smooth() functionality within the package ggplot2. A guideline through intercept = 0 is shown to highlight the discrepancy between the two parameters.

**Fig 5:**
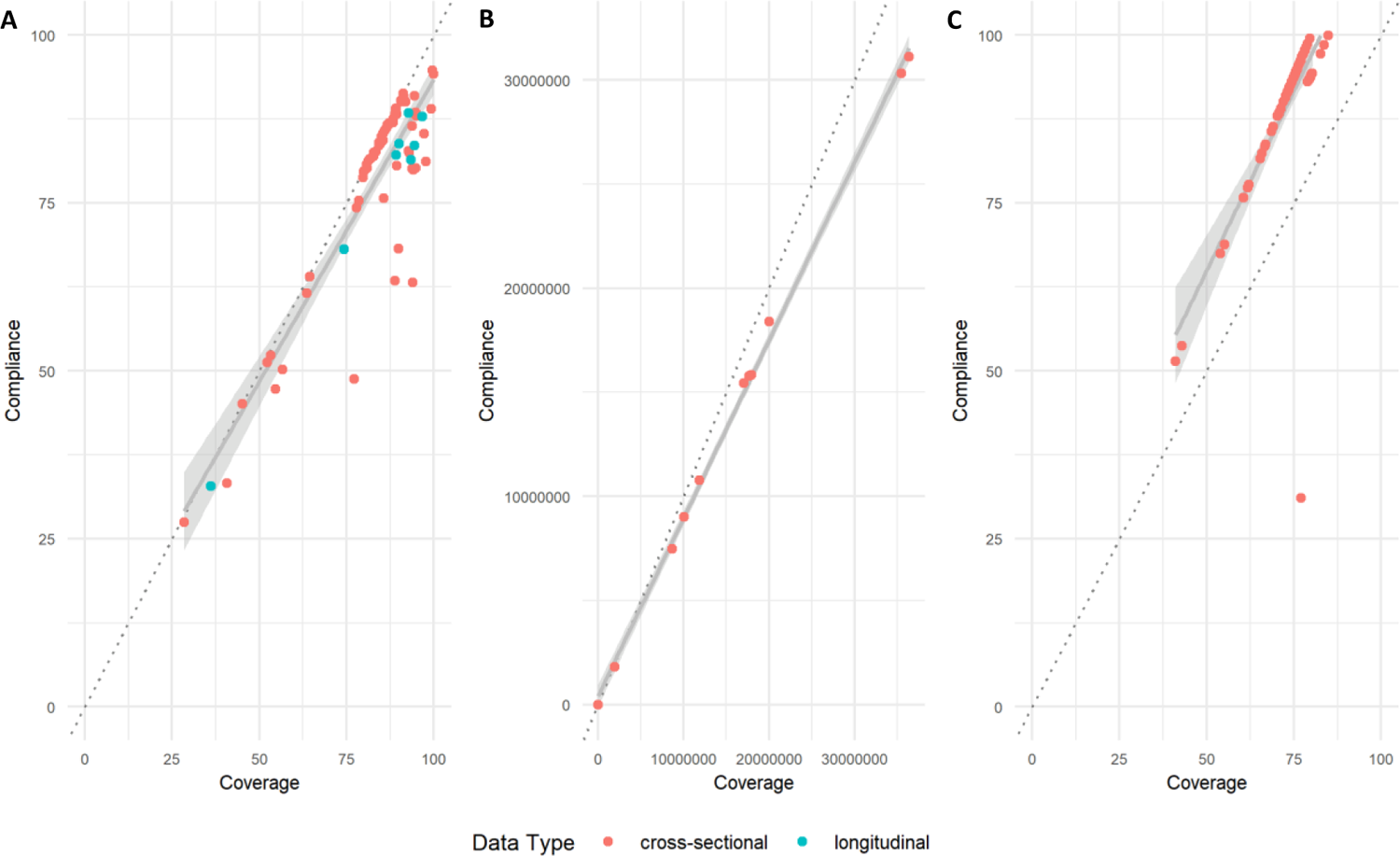
The variety of parameters used to describe cross-sectional and longitudinal compliance demonstrated, showing the most frequently used parameters both correctly and incorrectly used to describe compliance. The frequency of parameter use is proportional to the size of the circle. Compliance is used most frequently to describe longitudinal compliance (n = 8), whereas treatment coverage was most frequently used to describe cross-sectional compliance (n = 4). The number of circles represent the number of parameters given for compliance. Figure hand-drawn by authors.

### Stratification and analysis of compliance by demographic and behavioural factors

Beyond the greater understanding benefited from considering compliance over coverage on programme impact, stratifying the former by demographic and behaviour factors will provide further information to highlight target groups for enhanced sensitisation and or mobilisation. A total of 28 studies analysed key factors elucidating positive and negative associations influencing compliance in the studied population. Three main methods were employed for the analysis of compliance strata, mainly univariate, bivariate, and multivariate regression (presented as adjusted odds ratios (AOR)), Chi-squared analysis, and proportional breakdowns. Beyond the age, gender and location strata previously mentioned, further analyses were performed against enrolled/non-enrolled SAC, highest education level achieved of respondent, socioeconomic status, community type (rural/urban), marital status, occupation, ethnicity, religion, and length of stay in current village location.

Compliance increasing with age was noted by 12 studies. Specifically, compliance to treatment increasing with age was reported by Osei et al. (2022) (53) (AOR 4.23 (95% CI: 0.84-21.19), adults aged >58, compared to 18-27 year olds), and Chisha et al. (2020) (56) (AOR 1.45 (95% CI: 1.25-1.69), SAC aged 10–14, compared with to 5-9 year olds). Aza’ah et al (2020) (57) presented this phenomena via Chi-squared statistics, noting the proportion of age groups complying with MDA increased from 22.6% (95% CI: 16.3–30.4%) of 10-19 year olds, to 60.3% (95% CI: 53.1–67.1%) of participants aged ≥ 50 years (χ^2^ = 64.1, df = 3, P < 0.0001), as did Minetti et al. (2019) (38), who noted MDA participation increased with age (χ^2^ = 8.75, df = 1, p = 0.003). Mirroring this, Kifle and Nigatu (2021) (42) noted low compliance in young age groups, (AOR 0.68 (95% CI: 0.77-1.76) young adults aged 15-24 years, compared to >45 year olds). The variety of methods used to present age-related risk-factors with compliance prevents a subsequent meta-analysis to be conducted. Inconsistent conclusions were drawn for gender-related compliance behaviour. Statistical significance was noted for children enrolled in school, compared to those who were not (AOR = 20.90 (95% CI: 17.41-25.08) (56), AOR 2.29 (95% CI: 1.13–4.61) Asfaw et al. (2021) (58)).

Intuitively, consistent treatment behaviour to consecutive rounds of MDA was noted to decrease parasite prevalence. Six studies noted an increased odds of compliance based upon previous compliance, the treatment behaviour of systematic treatment. This phenomenon was presented with differing outlooks, varying between increased compliance based upon previous compliance (AOR 10.58 (95% CI: 5.78-19.38)) (53), willingness of future compliance by Tilahun and Fenta (2018) (34) (AOR 5.78 (95% CI:2.44–13.68)), and non-compliance related to previous non-compliance reported twice; (AOR 1.620 (95% CI: 1.051-2.497)) (38) and (AOR 9.18 (95% CI: 2.43-34.65)) by Dickson et al. (2021) (50). Campillo et al. (2021) (32) further developed the analysis, predicting the number of subsequent rounds required to clear treatment based upon current compliance to available rounds of MDA.

Aside from demographic factors, perceptions of the MDA activity and the CDD delivering the treatments were also considered by some studies. Common themes included awareness of target disease and the role of MDA, perceived susceptibility to target disease, CDD dedication, health benefits or adverse events related to drug ingestion. Prior knowledge of MDA was noted as significant by two studies, Asfaw *et al.* (2021) (58) noted a 69% lower compliance to MDA, between prior awareness of date versus date and location of MDA activity (AOR 0.31 (95% CI: 0.12–0.82)), whilst Krentel *et al.* (2016) (55) noted compliance increasing with prior awareness of MDA activity (AOR 2.59 (95% CI: 1.0-6.7)). The perception of drugs to be beneficial to participants was also noted to be significantly associated with increased compliance; (AOR 7.33 (95% CI: 4.13–13.02)) (34), (AOR 5.25 (95% CI: 2.55–10.82)) (53), and (AOR 10.74 (95% CI: 5.1-22.6)) (55). The perceived dedication and attitude of CDDs, as well as the protocols of height measurement were analysed by some authors, proving to be significant factors in increasing participant compliance. Travel time to the MDA distribution site was only collected by one study, delineating an individual’s will to take MDA, with the practicality of doing so (AOR 0.22 (95% CI: 0.14–0.35) >60 minutes, AOR 0.08 (95% CI: 0.04–0.17) 30–60 minutes), reference: < 30 minutes) (58). Participant comprehension of parasitic disease (AOR 13.68 (95% CI: 1.65–113.57)) (59), or the purpose of MDA (AOR 2.27 (95% CI: 1.44–3.59)) (58) was noted to be significant. Similarly, internal cues to action such as the assumption of own infection status, susceptibility of disease and fear of disease was shown to be significant (AOR 3.628 (95% CI: 1.96–6.73)) (60). It must be noted however, that despite these large AOR presented throughout the literature, the associated confidence intervals are very wide, reducing the reliability of this reported statistic.

## Discussion

This review highlights the paucity of individual longitudinal compliance studies available in the literature, despite publications dating back to 2004 (16), and more recently by Shuford *et al.* (2016), detailing the need for increased individual longitudinal cohort based compliance monitoring and reporting in the body of NTD research. The review aimed to compare reported individual longitudinal compliance recorded by CDD, referred to as DOT. The lack of publications resulted in the broadening of inclusion criteria to include all papers reporting compliance at all, analysing the few individual longitudinal studies, with longitudinal compliance self-reported by participants, and both repeated- and cross-sectional compliance studies.

This review also reports a large difference between coverage (receiving treatment) to compliance (swallowing treatment) when comparing the data reported (or aligning data reported to the definitions used in this review) by studies, mirroring the 22% coverage-compliance gap previously published by Babu *et al.* (2014) (14) and the 12.1% gap reported by Sitikantha *et al.* (2019) (61). In practice, this highlights the fact that a significant proportion of individuals that are un-treated are considered within the parameter of coverage. As such, programmes targeted with reaching coverage-based targets, conflating this parameter with compliance, could result in the premature cessation of programme efforts (55). For example, if coverage reaches a defined target e.g. 75% of SAC, and compliance is only 80%, then for a population of 1000, 750 will have received treatment, but only 600 will have swallowed treatment. This assumes the treatment has a 100% efficacy against the parasite, which is not always the case. For instance, albendazole is distributed in STH endemic areas, yet has a range of efficacies for different species, from *Ascaris lumbricoides* (91.4%), to *Trichuris trichiura* (50.0%) (62). As such, it may be demoralising when high coverage rates do not translate into efficient control with associated declines in infection prevalence and intensity. Beyond the clear definition of parameters used, a stipulation from WHO to routinely report compliance alongside coverage would be very beneficial. Control programmes targeted with reaching elimination of transmission that neglect to report compliance will be hindered in their monitoring accuracy if compliance is not recorded accurately.

Currently, there is heterogeneity between parameters and their definitions across species-specific guidelines provided by WHO. For example, in the case of lymphatic filariasis control, drug coverage is defined as the proportion of the *eligible* (also referred to as targeted) population that *swallow* drugs. However, in the same guideline, the programmatic drug coverage (defined as the proportion of the *total* population swallowing drugs) is recommended to reach 80%. Two different denominators are defined for the coverage parameter. Drug coverage surveys are also suggested whereby a sub-sample of the population are questioned on their treatment behaviour, including coverage and compliance treatment behaviours (63).This is the first instance of the notion to record compliance in NTD research. Concurrently, guidance provided for STH control defines coverage as the proportion of the target population *reached* (64). Systematic non-treatment (referred to as systematic non-compliance) is mentioned in the context of lymphatic filariasis control, defined as the individuals who never ingest the medicines in any MDA round, but is absent from the analogous STH control guide. It would be beneficial for future editions of the WHO roadmap, and species-specific control guidelines, to clearly delineate between coverage and compliance parameters, as current editions are non-comparable between species. These definitions need to be adopted by WHO across all NTD infections. The mis-match of parameter definitions currently provided by WHO will be particularly problematic for the species recently targeted for integrated control, as these will need to be adhered to across research groups and programmes for synchronous and efficient monitoring and evaluation. The recommendations provided by these guides resonate through the literature. As such, the continued misclassification of MDA parameter definitions in NTD research is highlighted by this review despite the advice presented by Shuford *et al.* (2016) six years previously (9). At a minimum, regardless of the parameter used by different studies and programmes, the calculation should be clearly defined within the methods enabling the scientific audience to interpret the data unambiguously. In the future, it is hoped that WHO guidelines will align on these parameters, setting the standard of clear definitions for the research community.

Since the publication of the last WHO NTD roadmap, considerable progress has been made globally in reducing the number of people infected with NTDs. As an increasing number of endemic communities are recording low prevalence, and the elimination of certain species as a public health problem has been achieved, completed by 17 countries for lymphatic filariasis and 10 countries for trachoma (1), a number of countries and regions can realistically look toward elimination of transmission. As previously noted by multiple authors, low parasite prevalence should first be validated with corresponding intensity measures, as commonly employed diagnostic techniques such as Kato Katz for the detection of intestinal nematode eggs in faeces show decreasing sensitivity in low prevalence settings, requiring more expensive, molecular methods (65). Furthermore, as is increasingly noted throughout the literature, to reach elimination of transmission, MDA delivery protocols would benefit from expanding school-based distribution to community-based distribution, as multiple studies have modelled and reported that only employing the former will leave pockets of infection in adults or preSAC who will continually reinfect the treated school aged children, resulting in the targeted communities to require indefinite treatment rounds as re-infection reverses treatment efforts (2,19,66). The coverage goal of this expanded protocol should be either increased to levels superseding that required for coverage to account for the reduced number of individuals swallowing treatment once reached, (the coverage-compliance gap), or include explicit compliance monitoring alongside coverage.

The importance of monitoring compliance at the individual level has been previously noted (18), and the impact of longitudinal compliance patterns upon control programme outcome is clear. Hardwick *et al.* (2021) created a framework of compliance (referred to as ‘adherence’), proposing the parameter *ωn* to measure the strength of association of previous compliance behaviour at MDA with that of a future round of MDA using stochastic individual-based simulations (7). This uses the notion of conditional probability, whereby the behaviour of an individual at one round of MDA is not independent of their behaviour at a previous round (systematic behaviour). Applying this framework to the age and gender-stratified compliance dataset collected by TUMIKIA across four biannual rounds of MDA, showed that despite high coverage figures, past behaviour-dependent compliance (or non-compliance) lowered the potential of hookworm elimination by 43% and 23% in two surveyed communities, respectively, compared to predictions based upon random compliance (behaviour at one round independent of behaviour at previous round) at each round of MDA. Past behaviour-dependent compliance was particularly apparent in males aged >30 years old. This conditional probability model demonstrates the impact that not only compliance, but demographic factors have upon reaching the goal of elimination of transmission. Of the 89 studies included in this review, compliance stratified by location, gender, and age were rarely provided. The importance of these descriptive strata is disproportional to their frequency. Presenting compliance data as a whole will potentially aggregate important behavioural heterogeneities within communities which need programmatic attention. Future WHO guidelines to Ministries of Health would benefit from not only including coverage stratifications by age and gender, but to reflect this in compliance reporting. MDA behavioural treatment patterns greatly impact MDA success, and thus will further impact the number of required rounds of MDA, and the magnitude of targeted coverage (to account for compliance behaviour).

NTD research is sometimes hindered by small budgets and limited study duration, thus the high cost of reporting infrastructure necessary for accurately monitoring individuals over several time points to capture longitudinal compliance may not be realistic. This is reflected in the proportion of longitudinally recorded compliance studies (n = 3) versus cross-sectional surveys using participant recollection (n = 22). As such, cross-sectional surveys conducted on a sub-sample of the treated population who are asked to recall if they have ever been treated, or preferably, how many treatment rounds they have participated in, is more realistic for most resource poor settings. Despite the decrease in accuracy of subject recall compared to DOT recorded by CDDs (which is also subject to its own biases), a cross-sectional snapshot of compliance in the population will provide a complementary measure to coverage and will help elucidate the impact of an MDA activity upon the local parasite burden. Furthermore, such programmes failing to reach targeted prevalence and intensity levels can accordingly target the demographic groups recorded with low compliance, as these fractions will arguably contain a high fraction of the total parasite population in a defined human community.

This review identified a single study reporting individual longitudinal compliance (20). This study also presented associations of key demographic and behaviour factors with compliance. Future studies and control programmes monitoring MDA activities, especially those in low prevalence settings, would be wise to follow this example. There are few such longitudinal studies currently underway which follow their study population at an individual level, namely the Geshiyaro project (67) and DeWorm3 (68), both following STH. These studies will have the potential to employ the recommended MDA monitoring strategies and provide individual longitudinal compliance data to the currently sparse pool of literature on the topic. Of most importance, is the inclusion of individual longitudinal compliance recorded by DOT. This is considered the gold standard of reporting in other disease fields that routinely follows longitudinal compliance such as HIV and TB treatment. Despite recently gaining attention in the NTD world, the monitoring of longitudinal compliance and treatment patterns across MDA rounds is well observed across other infectious and non-communicable diseases such as that to HIV treatment (69) and ACE inhibitors (70). The critical importance that systematic treatment patterns of longitudinal compliance impinges upon disease control has led to the development of Medication Event Monitoring System (MEMS), an electronic sensor embedded within medication bottle caps used to track compliance (71). This monitoring system is implemented for patients taking medication for a range of disease pathologies such as bipolar disease (72) and schizophrenia (73), to ulcerative colitis (74). Lessons learnt from other such areas of public health are of high relevance to NTD control programmes.

## Conclusions

There is a great lack of individual longitudinal compliance monitoring of MDA treatment in the NTD research landscape. This review has shown that there has been a slight, but steady, increase in the number of papers published which contain clearly defined compliance measurements, from a very low baseline. In the future, it is hoped that this trend continues, but at a much greater rate given the importance of these patterns to MDA impact on parasite prevalence and intensity. It must be noted, however, that the individual monitoring of compliance is financially and temporally expensive since it requires longitudinal cohort studies. In addition to enhanced accuracy of coverage and compliance recording required, a greater focus is also required on recording demographic variables such as population size and age structure. Ideally, a census or register of all participants must first be taken to monitor MDA behavioural patterns. As such, prospective cross-sectional studies interviewing participants on their treatment history at one point in time are proving to be the most popular approach. Despite this not being the gold standard of longitudinal compliance measurement, the information that a control programme or local ministry of health will garner, such as the definition of systematic or random compliance patterns in defined demographic sub-groups, will be crucial for guiding future MDA rounds to reach the goals of taking infection to very low levels or indeed eliminating transmission in defined populations or regions. Greater attention by WHO is required to unify definitions on both coverage and compliance, and guidance for the broad range of NTDs that are controlled by MDA programmes.

## Data Availability

A full data extraction file is available in S1 4.

## Acknowledgements

The Geshiyaro Project is funded by the Children’s Investment Fund Foundation (CIFF), grant number: R-1805-02741. The authors declare no competing interests.

## Supporting Information Captions

**Supporting Information 1:**
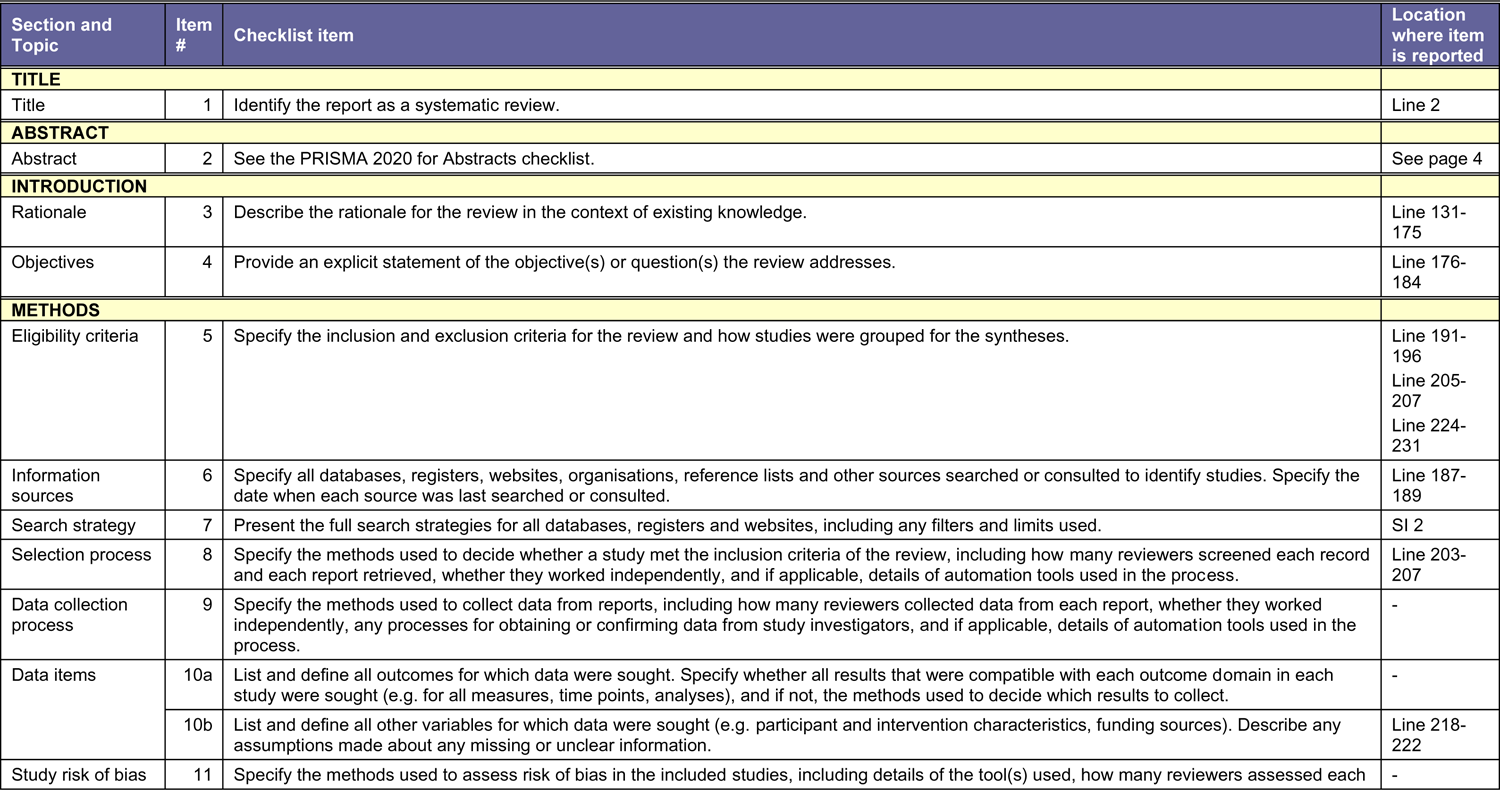
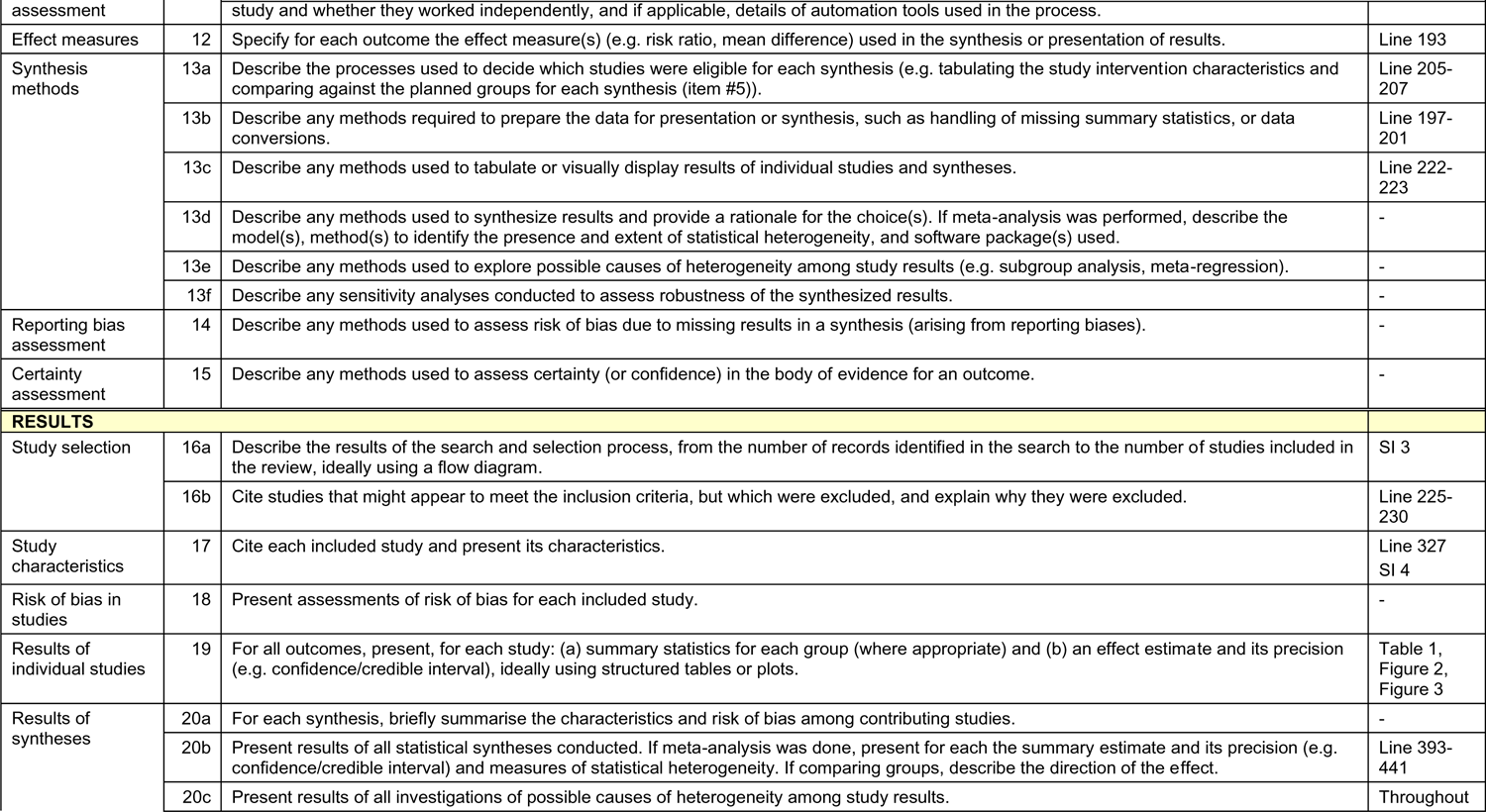
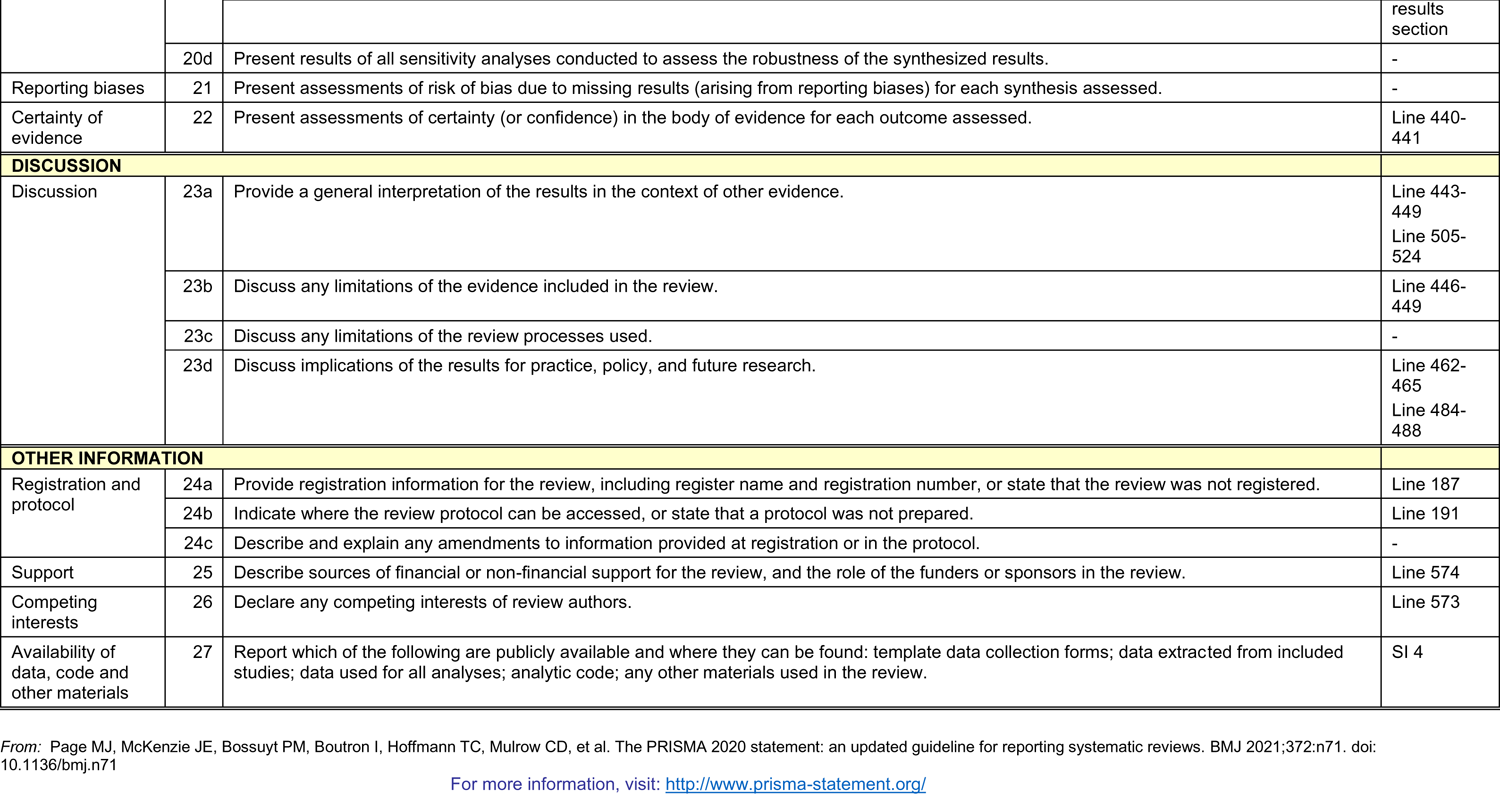
PRISMA guidelines for full text of review and abstract.

**Supporting Information 2:** The search strategy employed to generate the review.

| Section and Topic | Item # | Checklist item | Reported (Yes/No) |
| --- | --- | --- | --- |
| **TITLE** |  |  |  |
| Title | 1 | Identify the report as a systematic review. | Line 2 |
| **BACKGROUND** |  |  |  |
| Objectives | 2 | Provide an explicit statement of the main objective(s) or question(s) the review addresses. | Line 19 |
| **METHODS** |  |  |  |
| Eligibility criteria | 3 | Specify the inclusion and exclusion criteria for the review. | Line 25-26 |
| Information sources | 4 | Specify the information sources (e.g. databases, registers) used to identify studies and the date when each was last searched. | Line 23 |
| Risk of bias | 5 | Specify the methods used to assess risk of bias in the included studies. | - |
| Synthesis of results | 6 | Specify the methods used to present and synthesise results. | Line 26-27 |
| **RESULTS** |  |  |  |
| Included studies | 7 | Give the total number of included studies and participants and summarise relevant characteristics of studies. | Line 27-28 |
| Synthesis of results | 8 | Present results for main outcomes, preferably indicating the number of included studies and participants for each. If meta-analysis was done, report the summary estimate and confidence/credible interval. If comparing groups, indicate the direction of the effect (i.e. which group is favoured). | Line 28-29 |
| **DISCUSSION** |  |  |  |
| Limitations of evidence | 9 | Provide a brief summary of the limitations of the evidence included in the review (e.g. study risk of bias, inconsistency and imprecision). | Line 29 |
| Interpretation | 10 | Provide a general interpretation of the results and important implications. | Line 29-32 |
| **OTHER** |  |  |  |
| Funding | 11 | Specify the primary source of funding for the review. | - |
| Registration | 12 | Provide the register name and registration number. | Line 24 |

From: Page MJ, McKenzie JE, Bossuyt PM, Boutron I, Hoffmann TC, Mulrow CD, et al. The PRISMA 2020 statement: an updated guideline for reporting systematic reviews. BMJ 2021;372:n71. doi: 10.1136/bmj.n71
For more information, visit: http://www.prisma-statement.org/

**Supporting Information 3:**
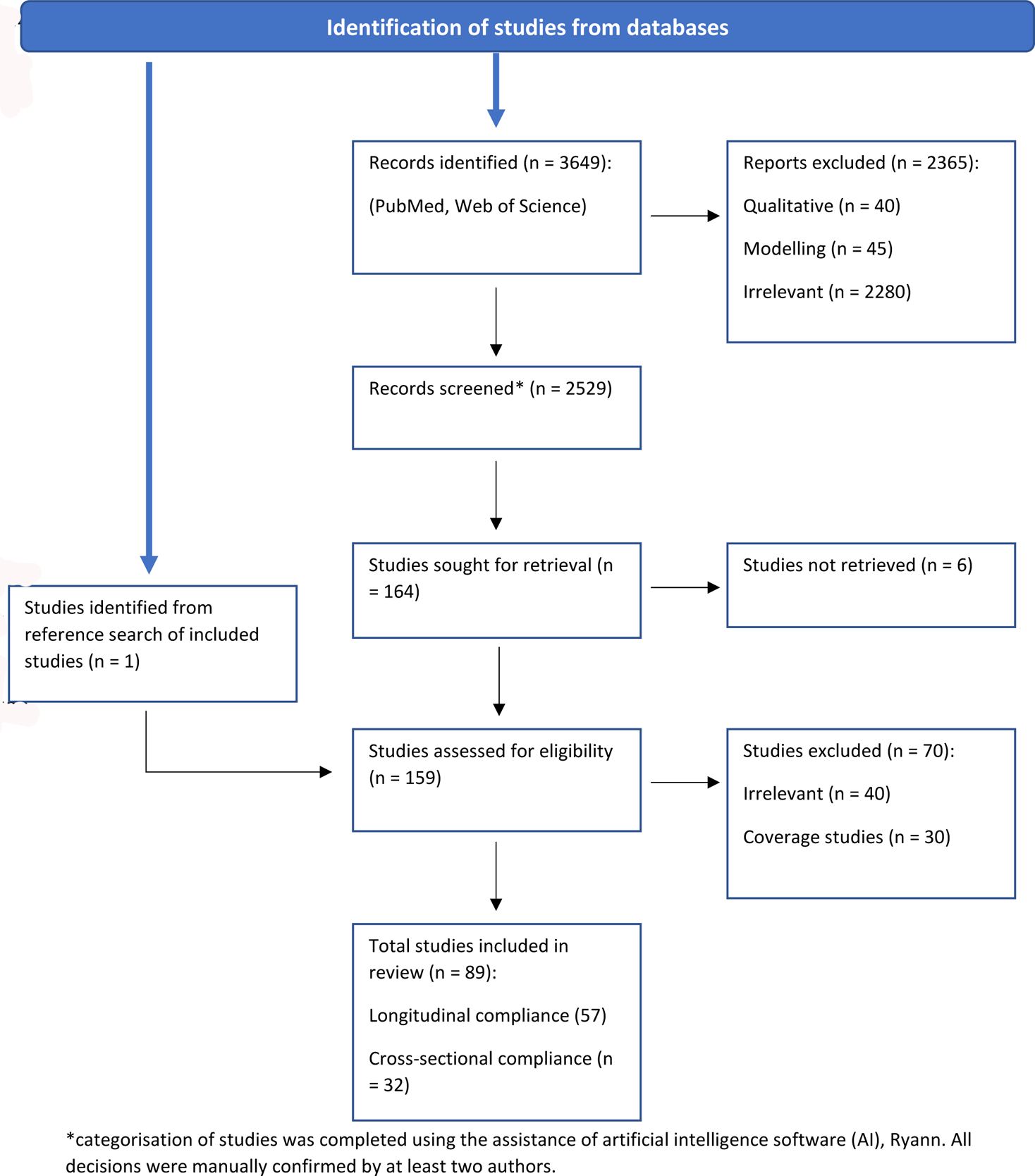
Workflow of screening and inclusion of studies as defined by the PRISMA guidelines.

**Supporting Information 4:**
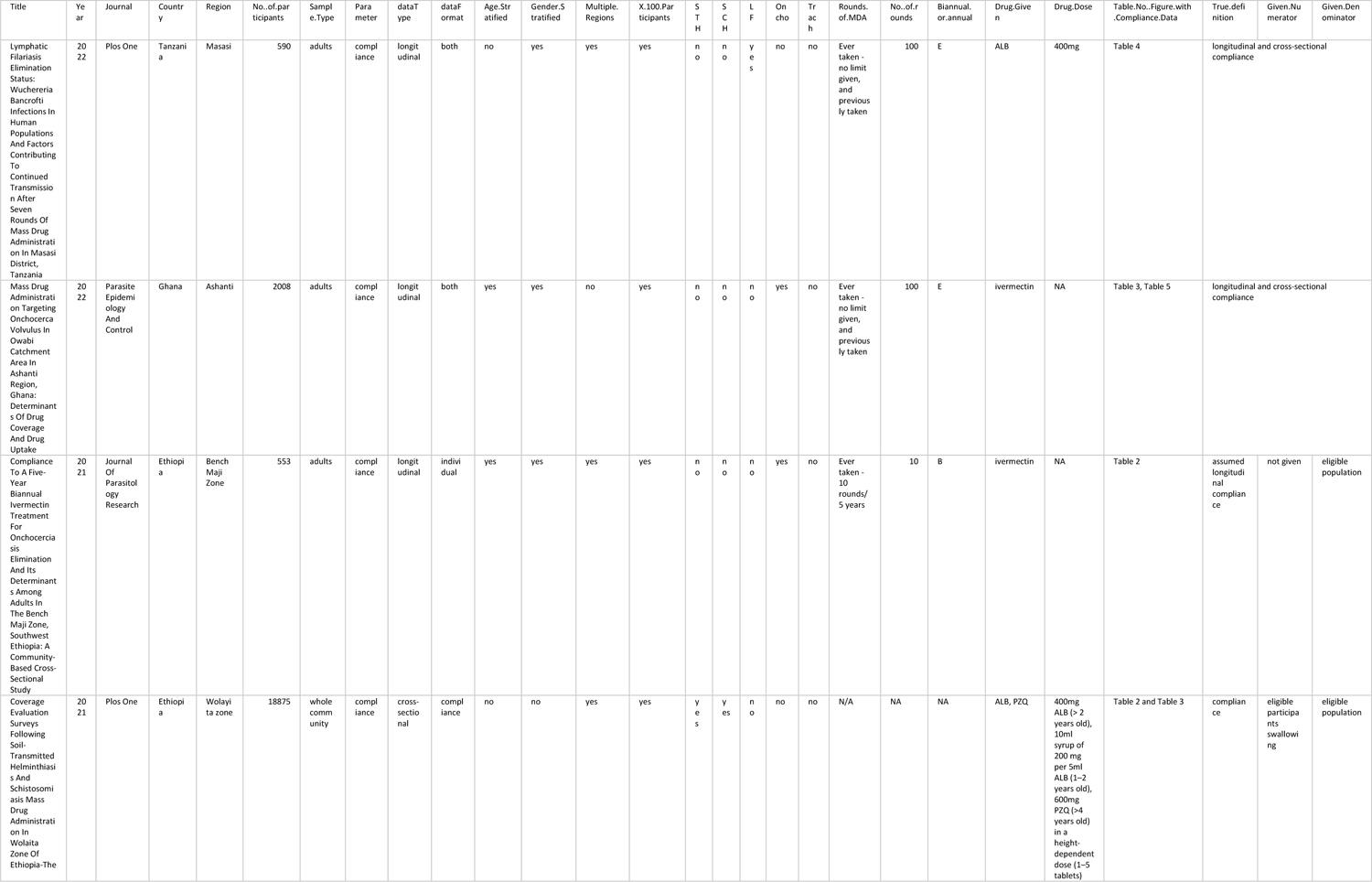
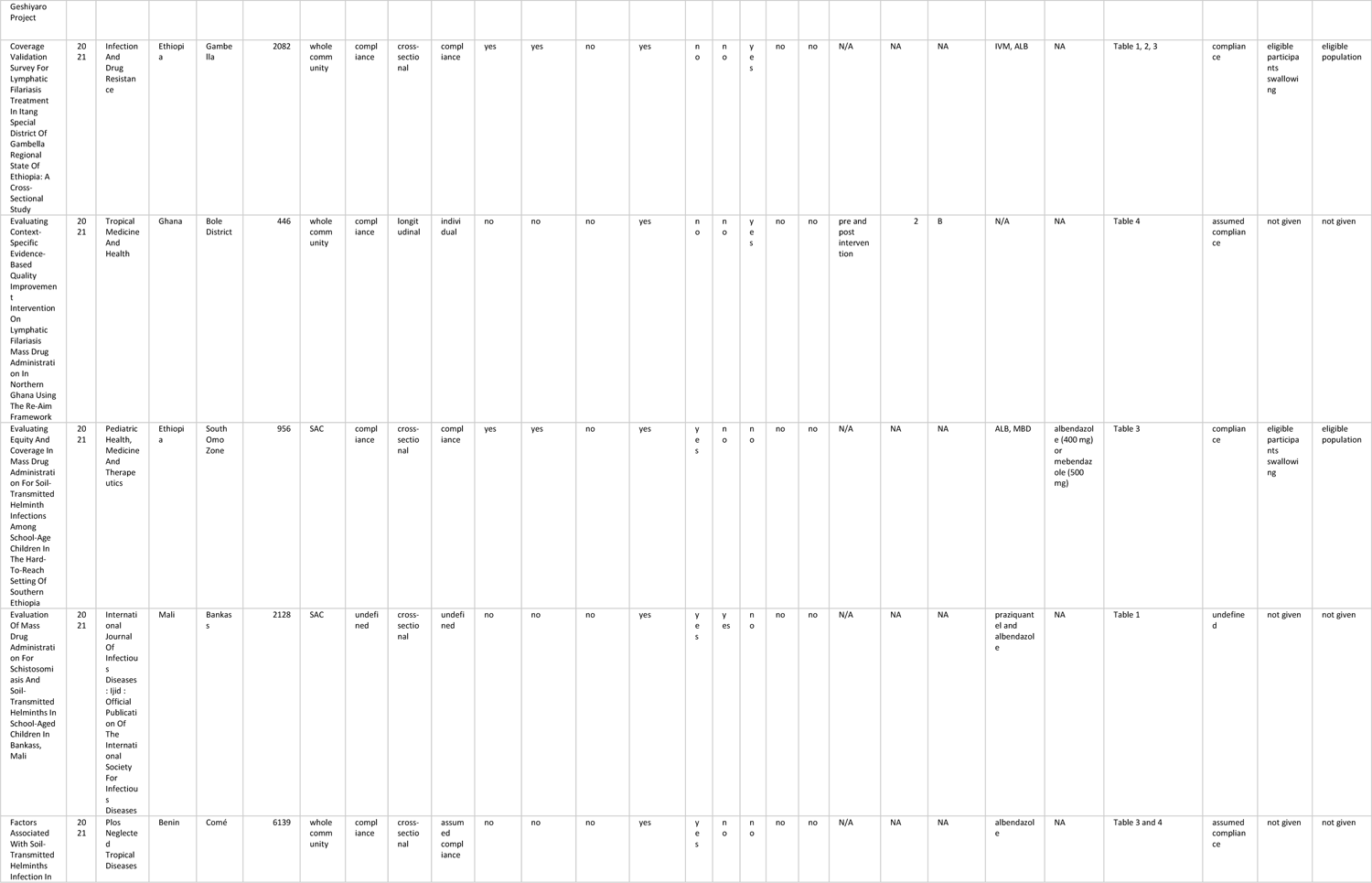
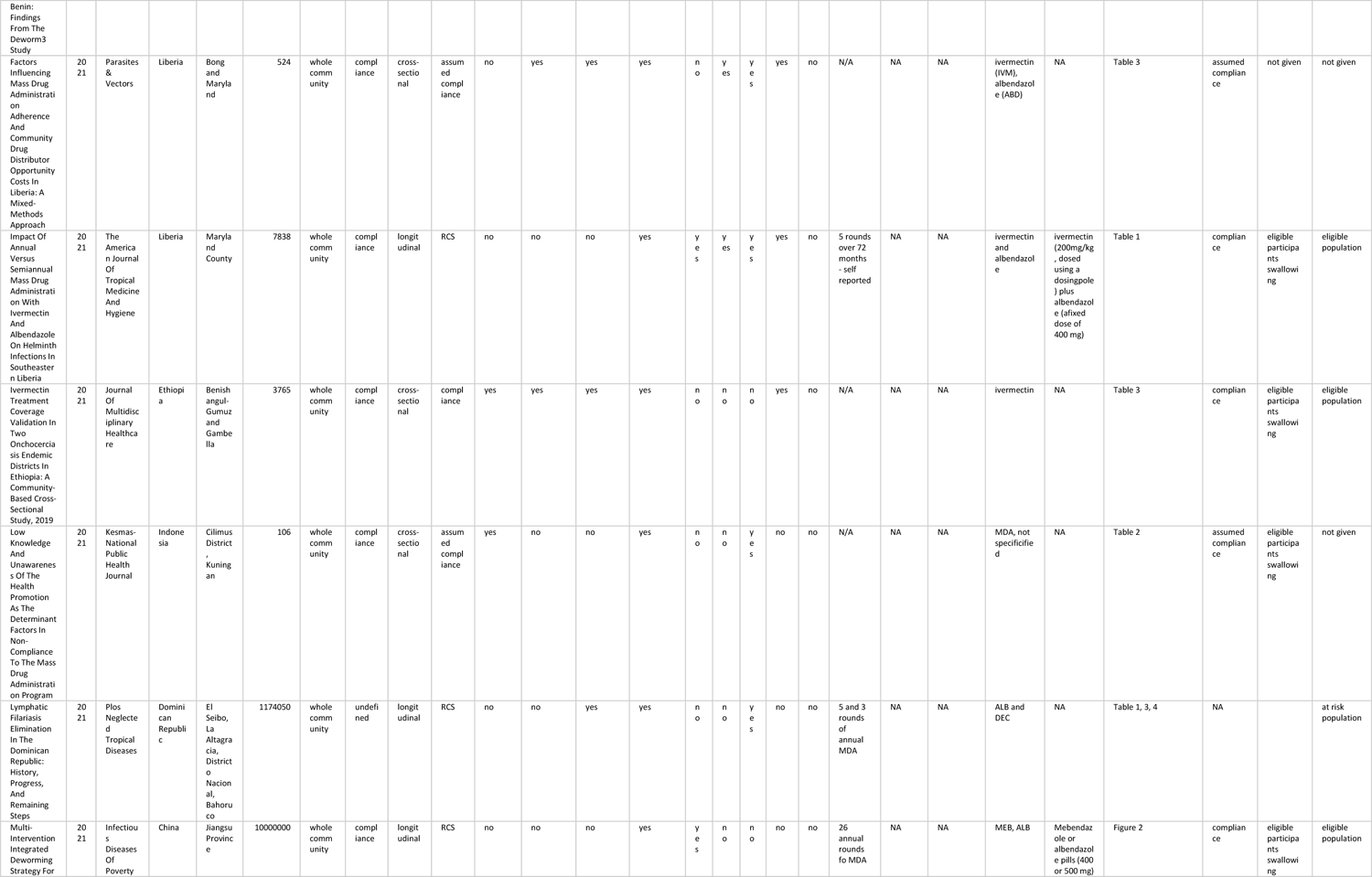
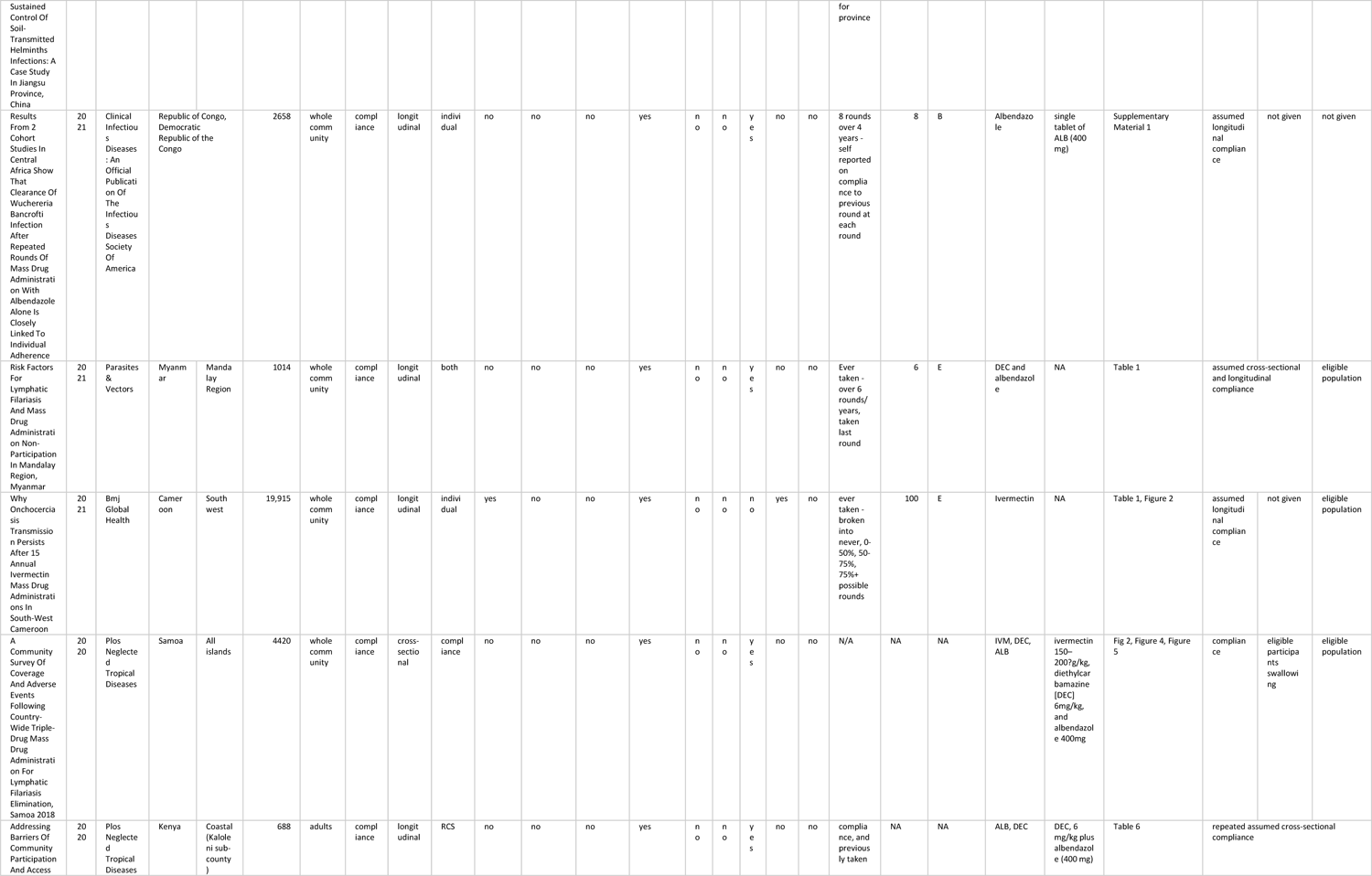
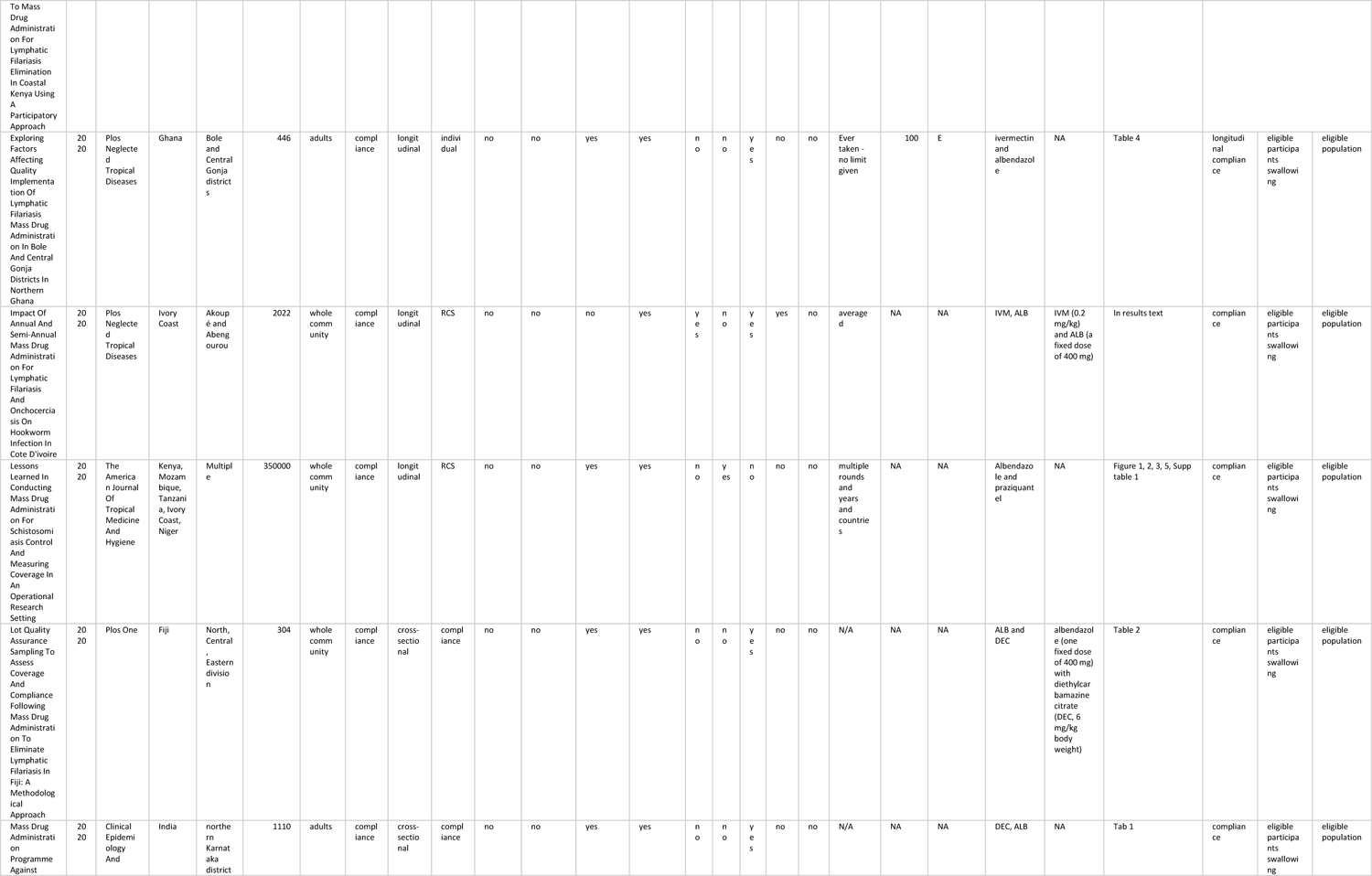
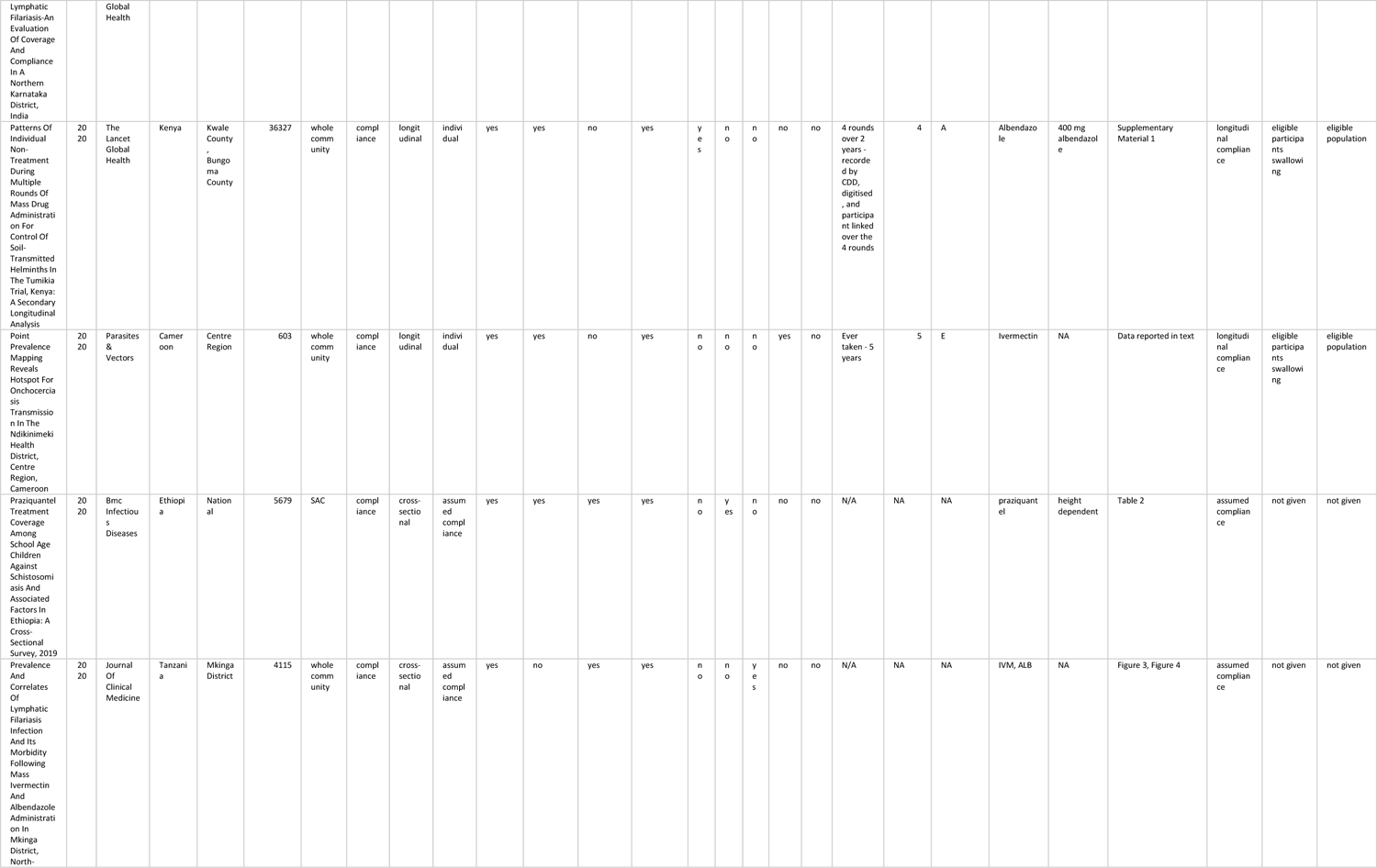
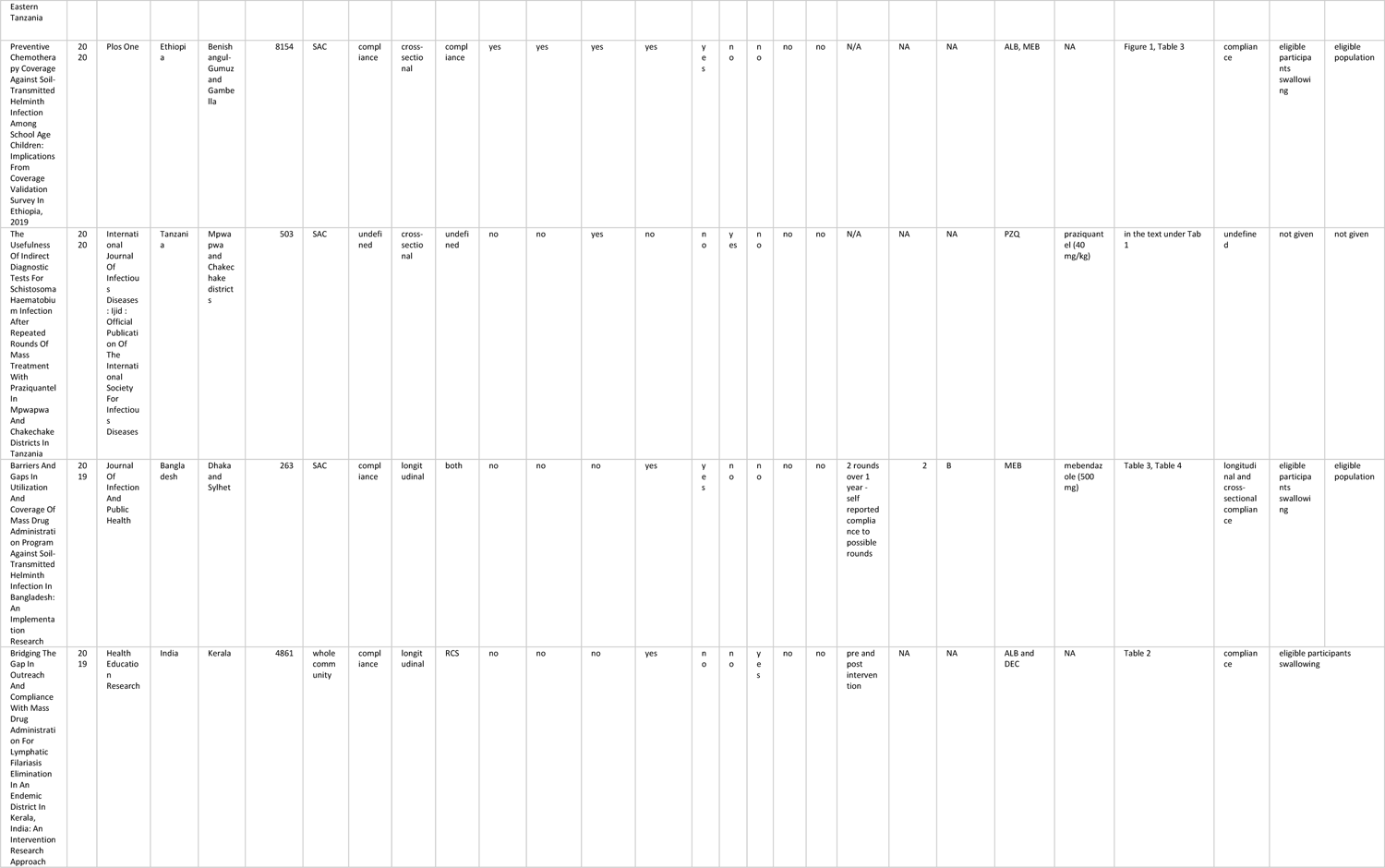
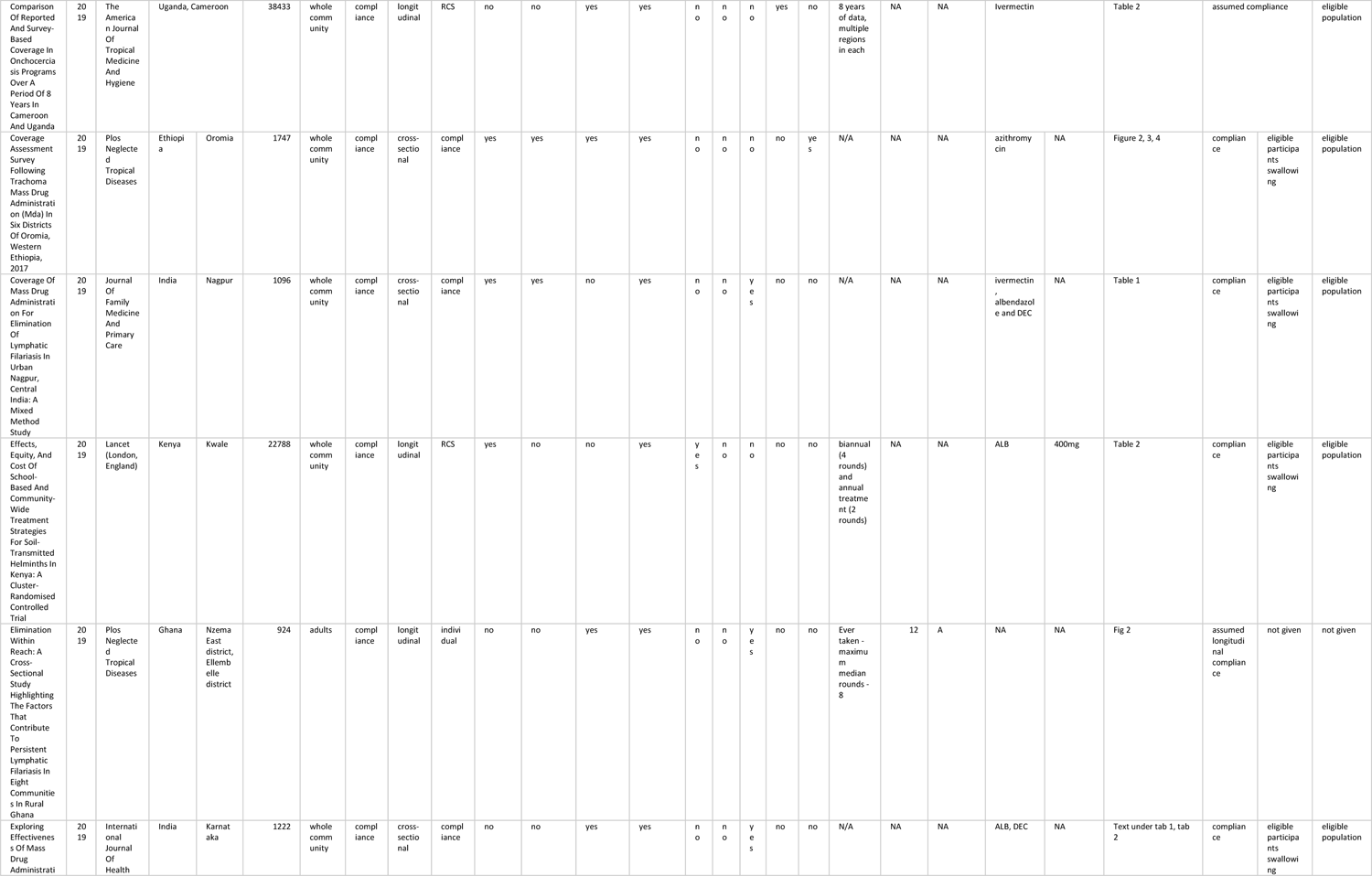
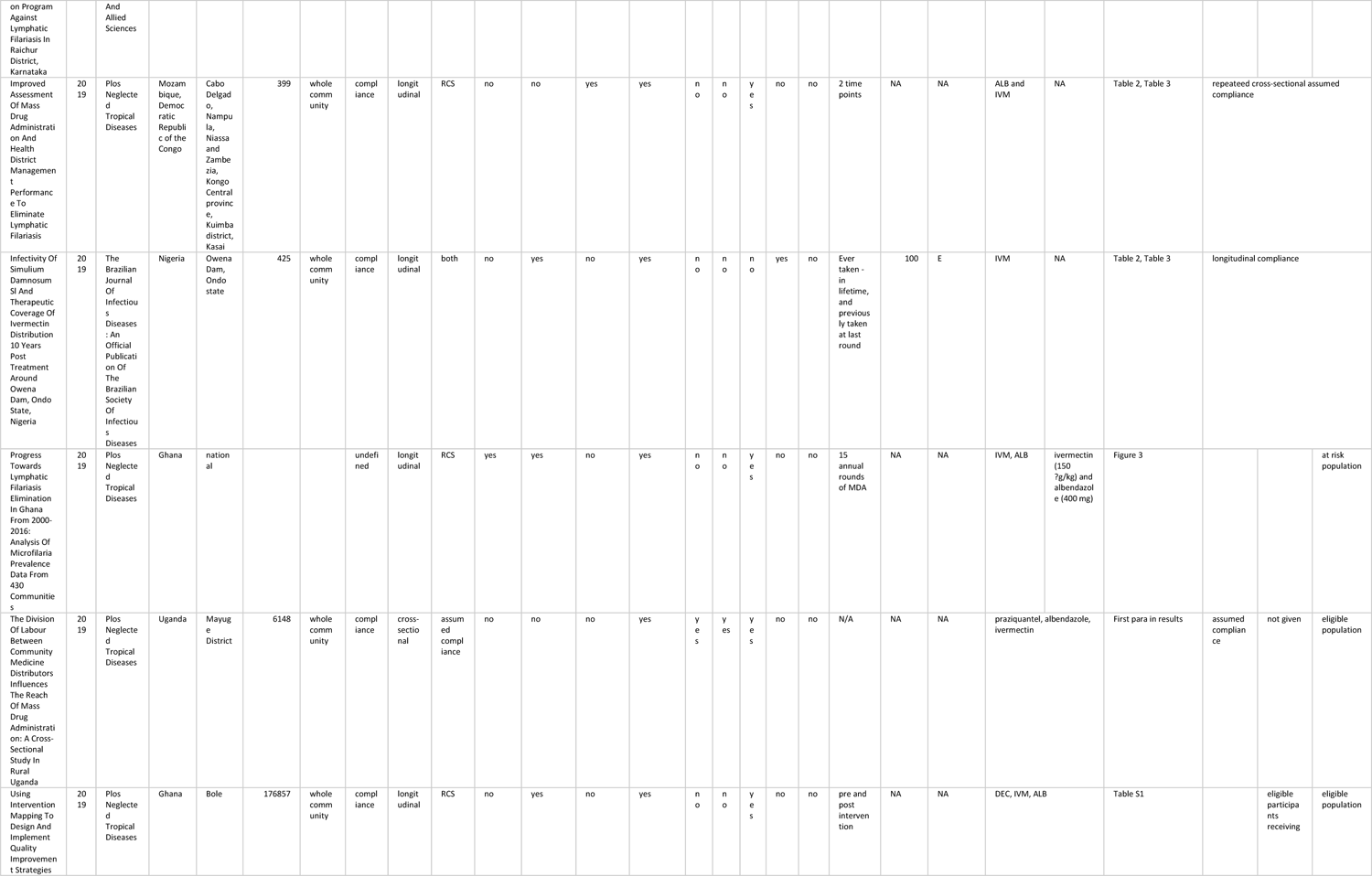
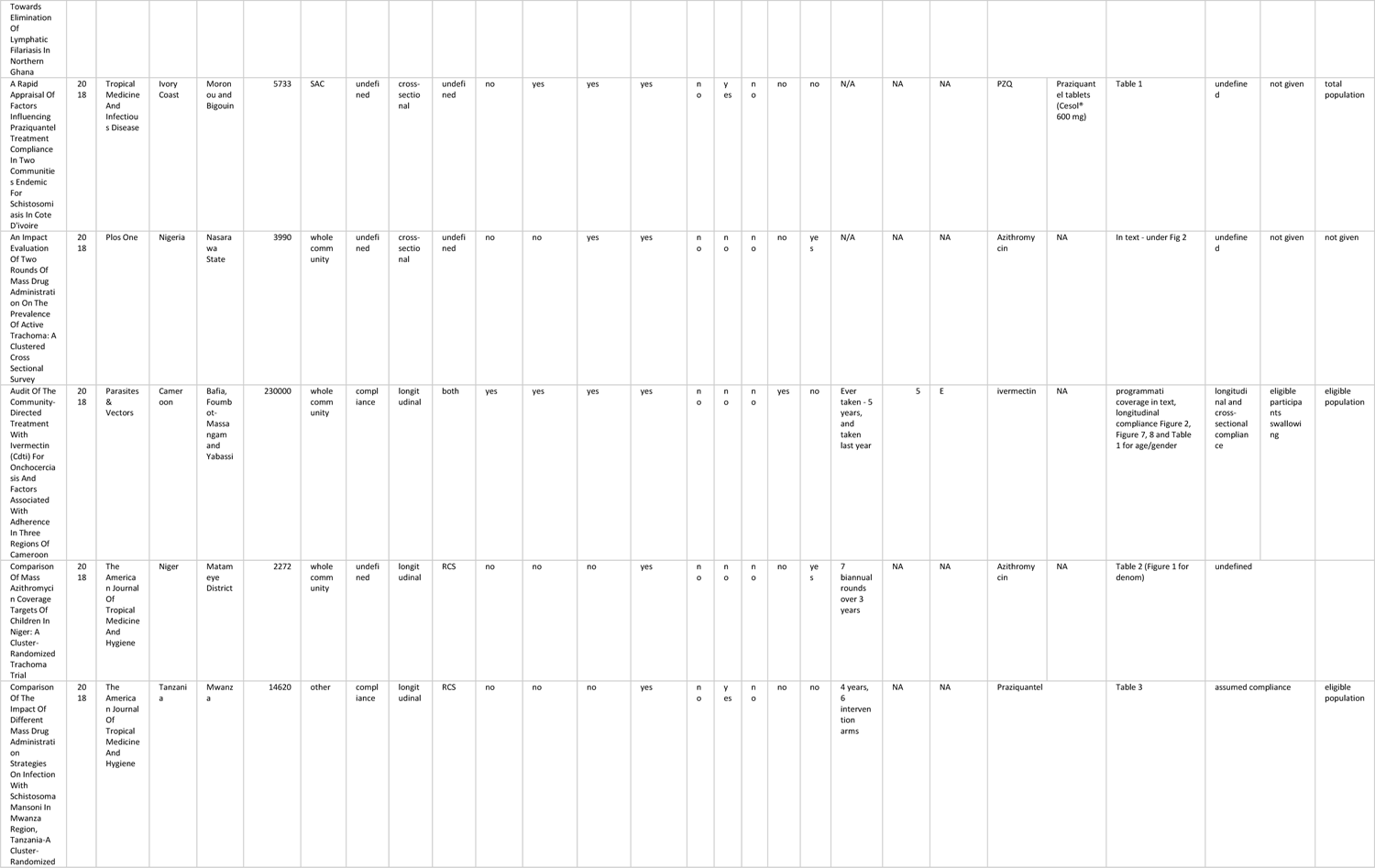
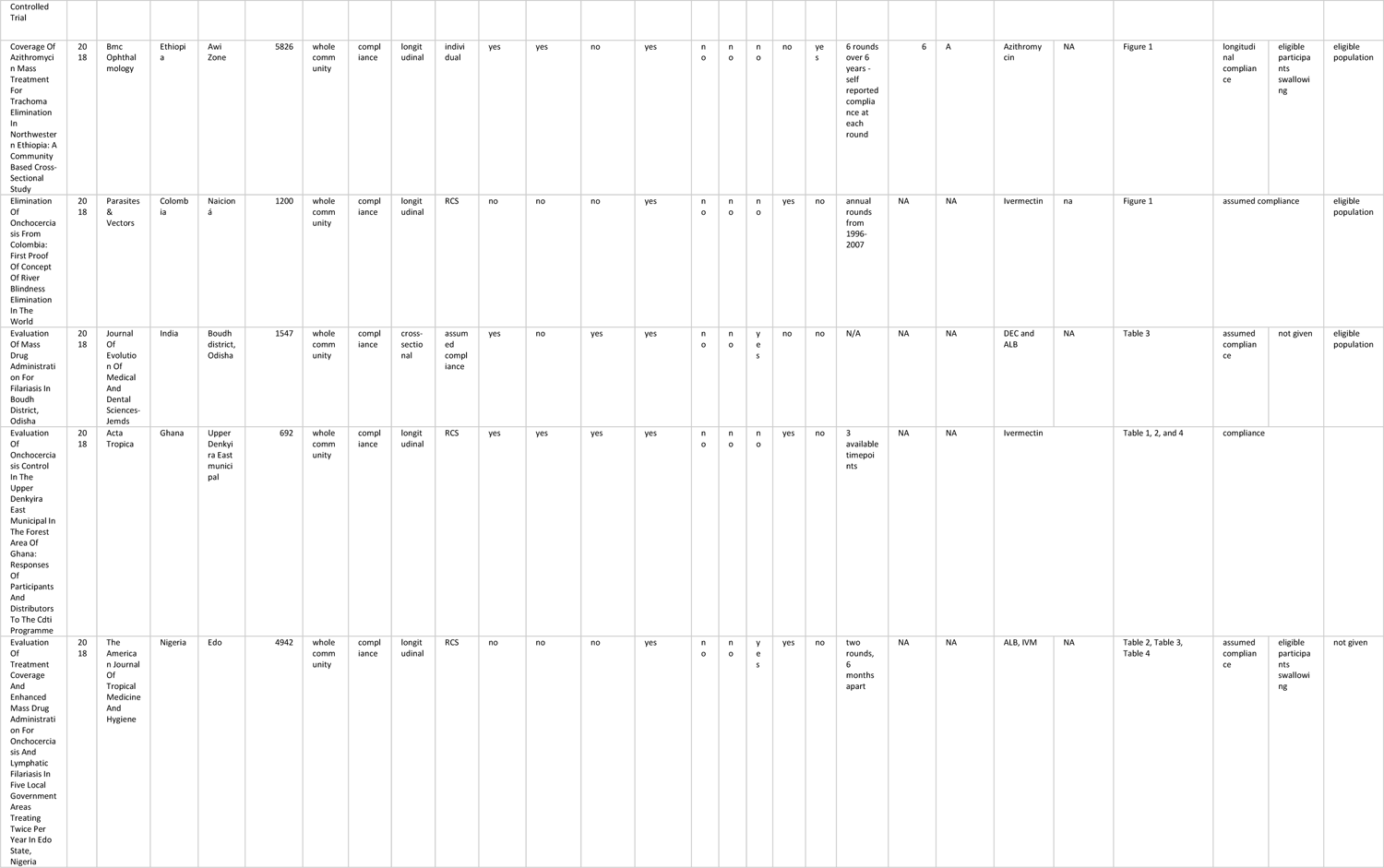
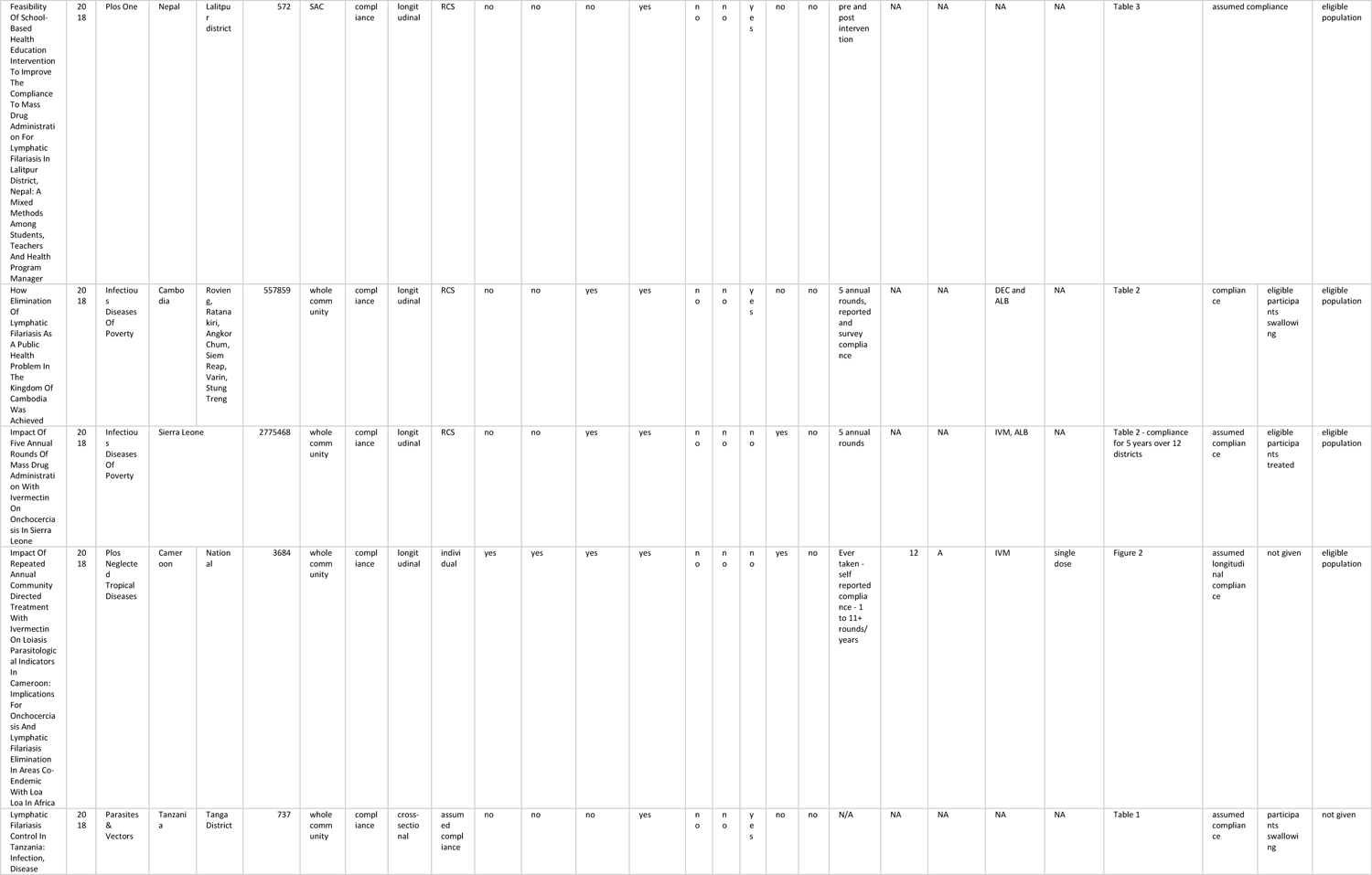
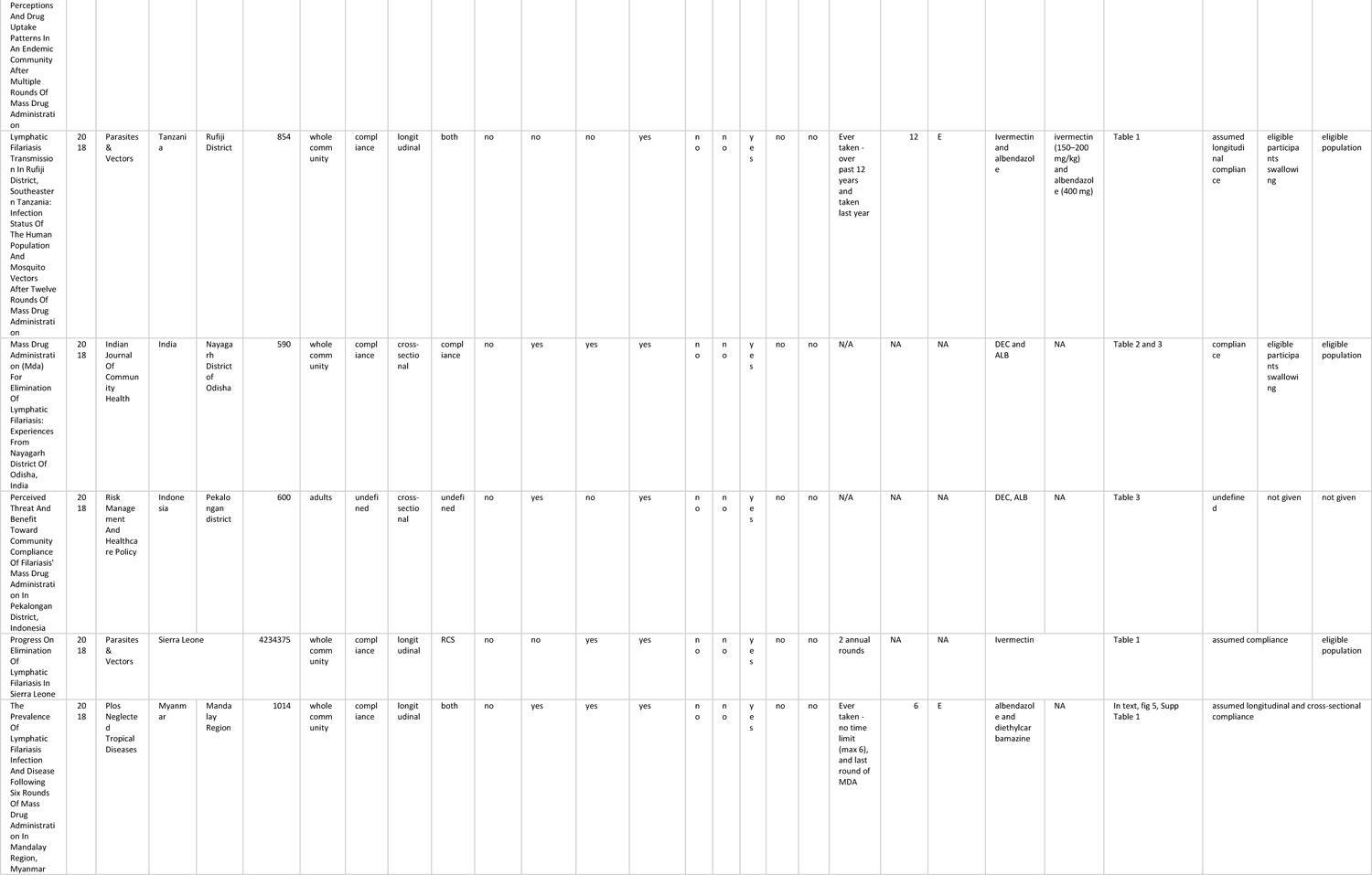
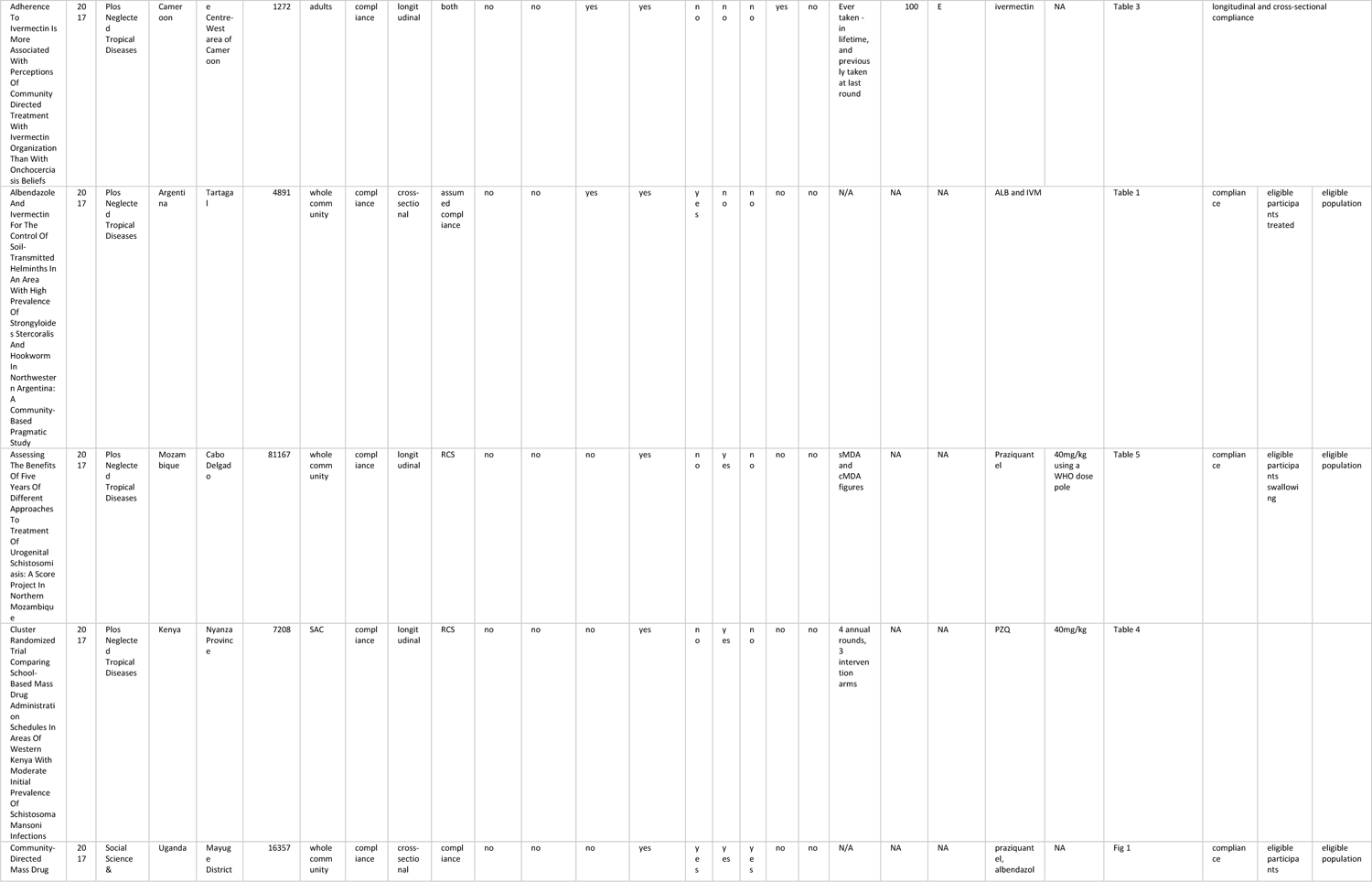
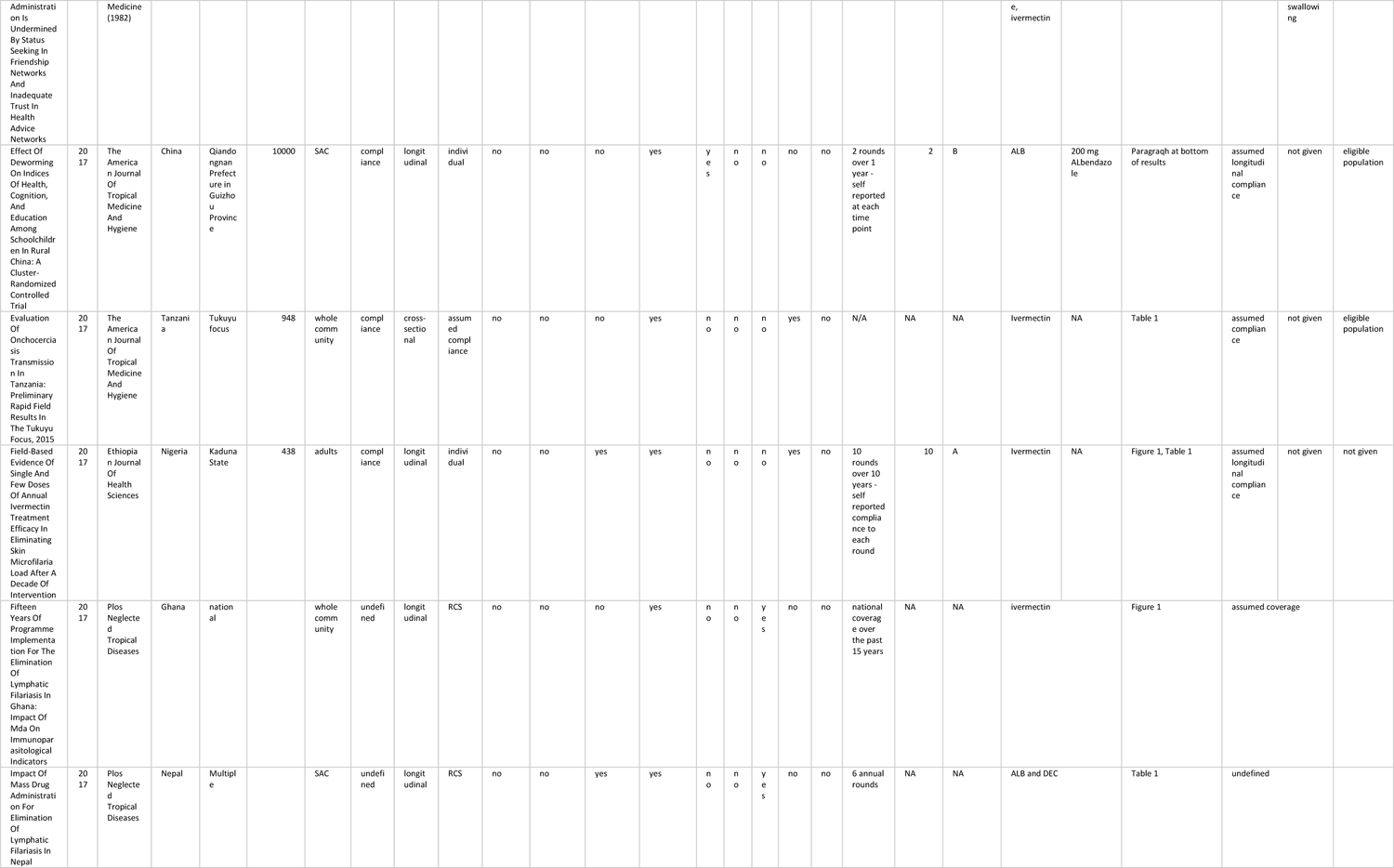
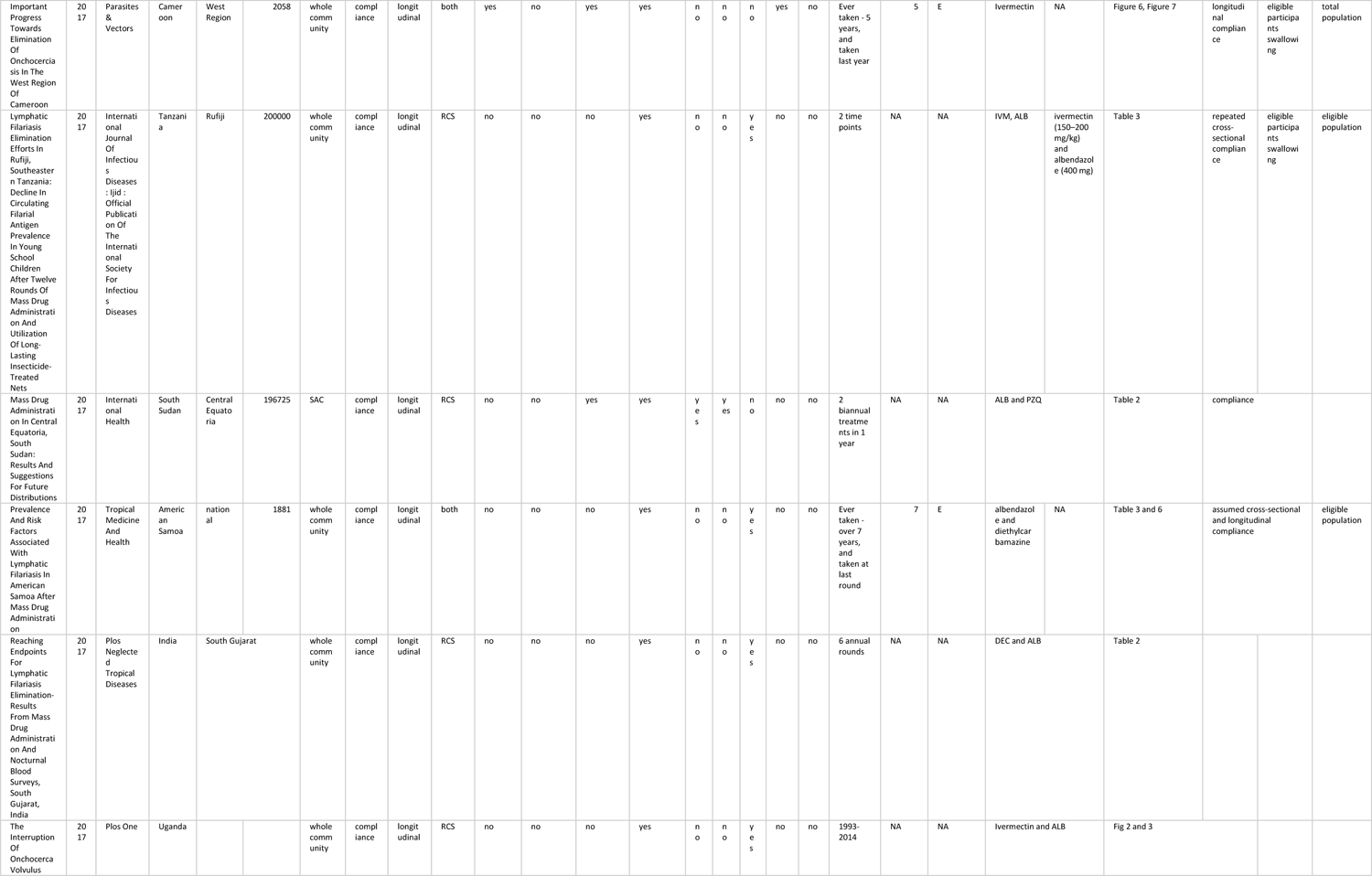
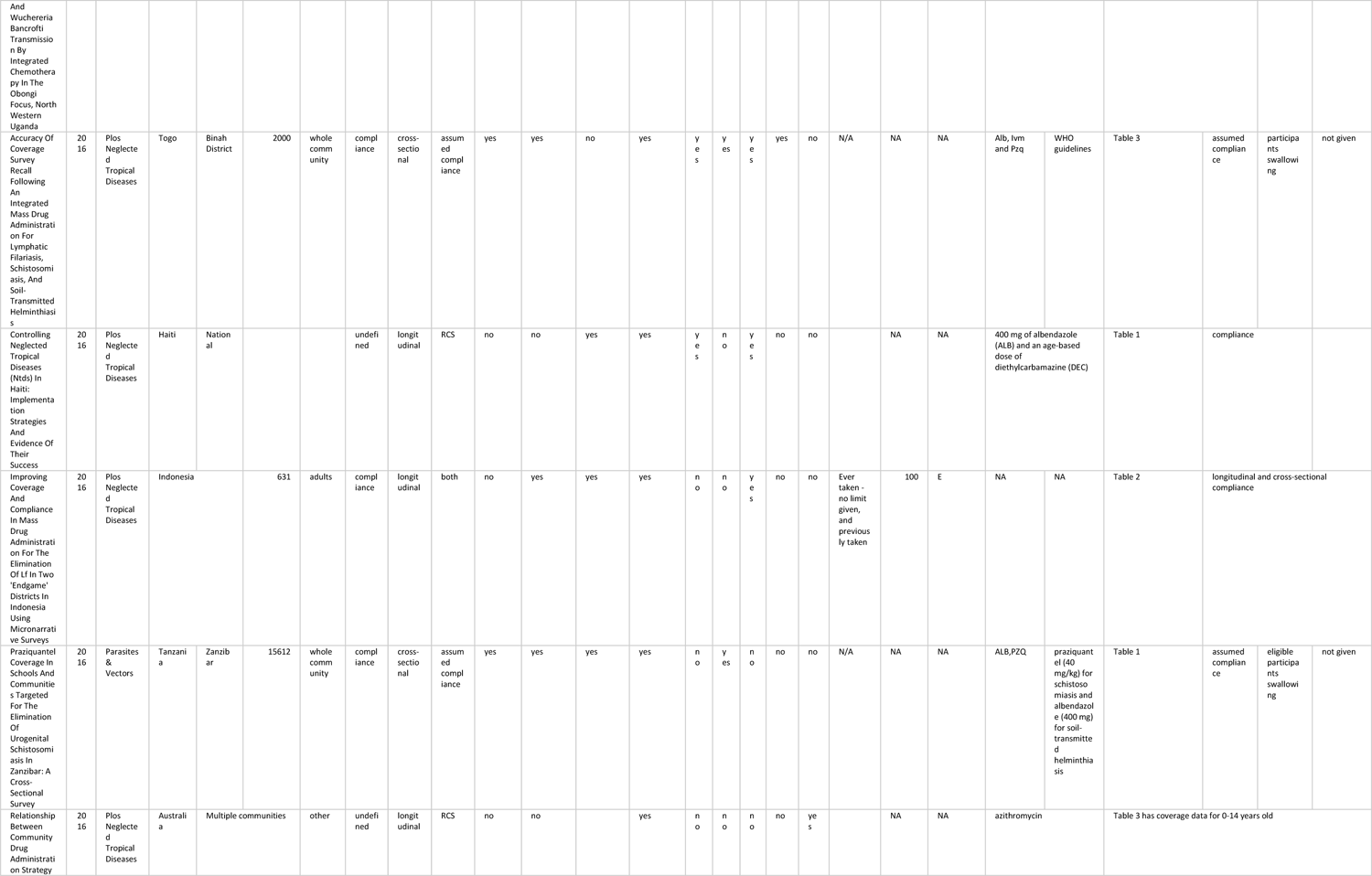
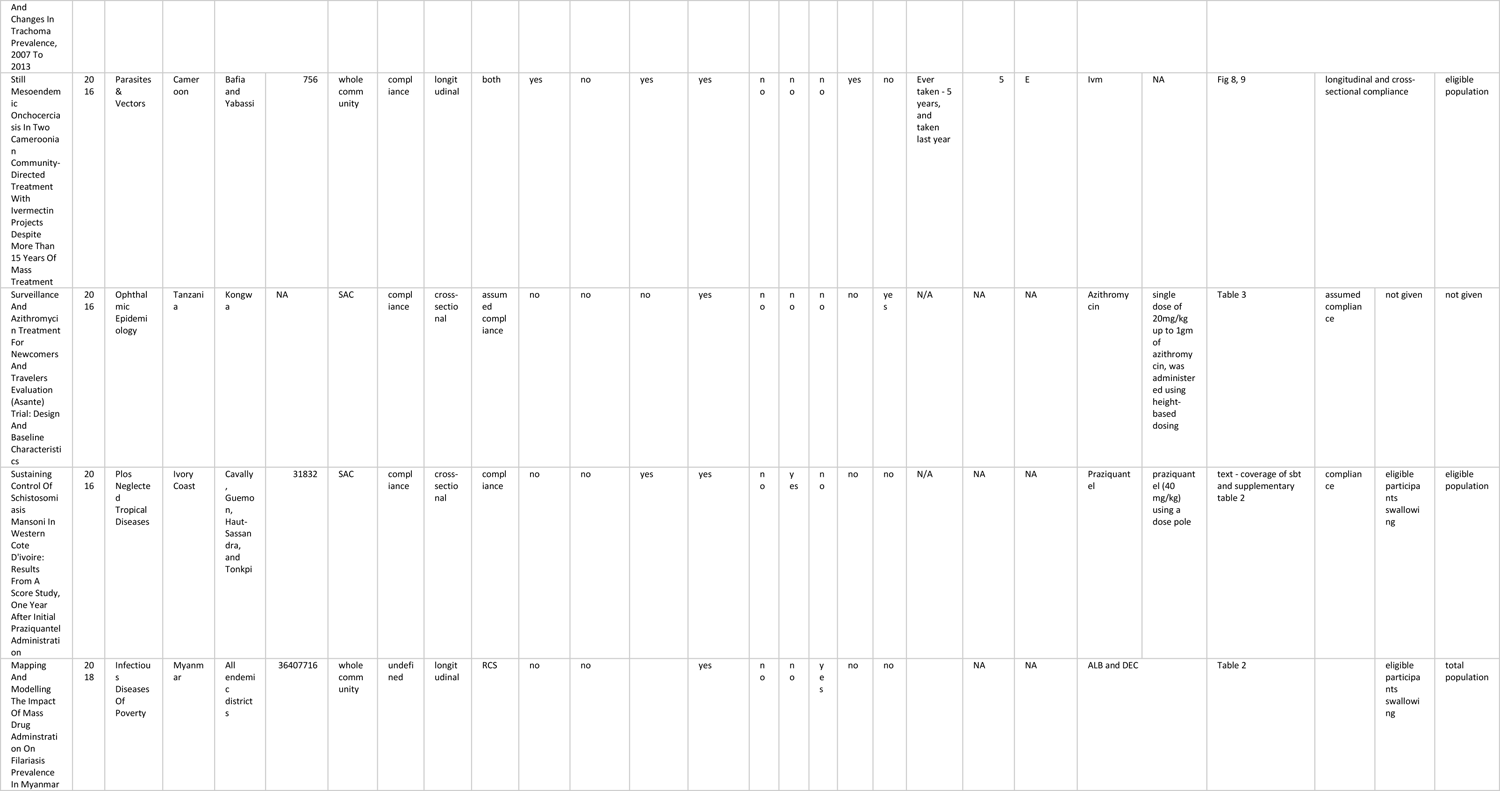
The complete 89 studies identified as containing compliance data.

